# Evaluating uncertainty for detecting performance loss and guiding deferral after model transfer between hospital systems

**DOI:** 10.64898/2026.11.03.26367064

**Authors:** Stefan Behling, Matthias Schulte-Althoff, Stefan Haufe, Elias Grünewald, Felix Balzer, Nicolas Frey, Sebastian Daniel Boie

## Abstract

Clinical prediction models often perform worse after transfer between hospital systems, but deterioration becomes apparent only once outcomes are known. We investigated whether uncertainty measures available at prediction time warn of performance loss and can help identify individual predictions requiring review. Three neural network architectures were trained on MIMIC-IV for in-hospital mortality prediction and evaluated on eICU-CRD. Measures included predictive entropy, mutual information from Monte Carlo dropout and deep ensembles, and representation density. After transfer, AUROC fell from 0.86–0.87 to 0.76–0.77 and calibration worsened, while entropy and mutual information were lower. None of the measures reliably indicated the reduced performance. Deferring the 20% with the highest entropy reduced the remaining error rate from 17.5–20.1% to 6.3–8.0%; the deferred patients included 91–99% of false alarms but only 2–7% of missed deaths, which carried low uncertainty. Uncertainty therefore supported selective review but failed to reveal performance loss.

## Introduction

Clinical prediction models, based on multimodal data, are increasingly deployed to detect deterioration and personalize treatment decisions^1–3^. Sequential deep learning models, including recurrent neural networks and transformers, have shown strong performance in intensive care time-series prediction^4,5^. However, very few medical providers can train their own models tailored to their patient population and medical practice. Increasingly, providers will deploy prediction models locally that are trained elsewhere. It is well-known that models developed using data from a single institution or health system may not generalize to other settings, and external validation remains uncommon^4^. Differences in patient populations, clinical practice, and measurement processes can compromise discrimination and calibration when models are transferred between institutions^5,6^. This deterioration may not be adequately reflected in model confidence^7,8^.

Consider a mortality prediction model trained in one hospital and used in another: it may return an estimate of 4% for a certain patient whose actual risk is several times higher. Nothing in the output signals that this estimate deserves less trust than one at the development site. Such failures can go unnoticed in routine care. A widely implemented proprietary sepsis model reached an AUROC of 0.63 in independent external validation, compared with 0.76 to 0.83 reported by its developer. It missed two thirds of patients with sepsis while alerting on 18% of all inpatients, and the shortfall was detected only retrospectively, from outcomes, after the model had entered routine use^9^.

Explainable AI (XAI) is a widely studied approach helping clinicians judge whether a prediction can be relied on^10^, yet clinical XAI evaluations rarely test whether explanations help clinicians detect incorrect predictions^11^. Explanations aim to show which inputs drove a prediction, not whether it is likely to be correct, and they can appear plausible for incorrect outputs^12^. Uncertainty quantification offers an alternative approach: it communicates alongside each prediction how cautiously the risk estimate should be treated^13^ and may flag unfamiliar inputs, potentially incorrect predictions, or cases that warrant human review^14,15^. Such measures may matter most where clinicians’ own error detection is weakest. Whether a given uncertainty measure actually flags incorrect predictions, however, has to be established with labeled data, as for calibration, and may depend on the data and the type of distribution shift. For in-hospital mortality prediction, evaluation of discrimination and calibration requires outcomes that become available only after predictions have been made, introducing a significant time lag between deployment and model validation. Uncertainty measures, by contrast, can be computed at the time of prediction, before the outcome is known^14^. By identifying outputs that require closer scrutiny, uncertainty quantification may complement model interpretation and support human oversight. It may also complement outcome-based evaluation within post-market monitoring frameworks for high-risk AI systems under the EU AI Act and for medical devices under the Medical Device Regulation^16,17^.

While our framework applies to all prediction tasks based on clinical routine data, we focus on in-hospital mortality as an example. Loss of performance after transfer has been reported across clinical prediction tasks^18^, and mortality prediction is no exception: ICU models developed at one institution lose discrimination and calibration at others^4,5^, as do established severity scores such as APACHE and SAPS, which require recalibration outside their development populations^19^.

In this article, we consider uncertainty measures that may indicate performance deterioration before outcome labels are available and we evaluate them for three evaluation levels. Cohort separation asks whether a measure distinguishes external from development cohort patients; error detection asks whether it ranks incorrect above correct predictions within the target cohort, and selective prediction asks whether deferring high uncertainty cases – withholding the model’s prediction and referring the patient to clinical review – reduces error among retained predictions. Previous medical-AI studies have examined these objectives in different settings, including out-of-distribution detection in clinical tabular data^20^, failure detection in medical imaging^21^, and selective prediction in multimodal intensive care data^22^. In ICU mortality prediction, uncertainty methods have been evaluated on artificially altered patient records from MIMIC-III^23^ and for sepsis prediction across MIMIC-IV, eICU-CRD and HiRID, strategies for adapting deep learning models to a new site have been compared systematically^24^.

Moreover, uncertainty estimates can deteriorate under dataset shift^7^, and on medical tabular data neither uncertainty estimates^25^ nor dedicated out-of-distribution detectors^20^ have reliably identified patients from a shifted population. Specifically, we evaluate output-, sampling-, and representation-based uncertainty measures for in-hospital mortality prediction under transfer from MIMIC-IV to eICU-CRD. After quantifying changes in discrimination and calibration, we assess each measure for (1) cohort separation, (2) prediction-error detection in the external cohort, and (3) selective prediction through uncertainty-based deferral.

We investigate the following research questions:

1) Do uncertainty measures increase after transfer to another hospital system, as a warning of deteriorating performance would require?

2) Do uncertainty measures identify the individual predictions that are likely to be wrong?

3) When human resources allow for manual review of a limited share of predictions, can the above measures suggest which patients to review to lower error rates?

The first question asks whether a measure registers that the patient population has changed; the second and third ask whether it registers that an individual prediction is unreliable. These are different properties, and a measure can have one without the other. Each question corresponds to one evaluation level described in the Methods: cohort separation, error detection, and selective prediction.

## Results

We report three base architectures (LSTM, GRU and transformer); each trained with ten random seeds. Values are means over the ten seed models; brackets give the 95% t-interval over seeds. Where two cohorts are compared, the difference was computed within each seed before averaging, so that the interval describes the paired difference across training runs.

### Cohort characteristics

The internal test set comprised 7,298 ICU stays from MIMIC-IV with an in-hospital mortality of 13.8%, and the external cohort 66,934 stays from eICU-CRD with a mortality of 10.9% (Table 1; Table 2). The external cohort was similar to the internal test set in age (median 66 vs 67 years) and sex (45.9% vs 44.6% women) but had a lower in-hospital mortality (10.9% vs 13.8%).

**Table 1.**
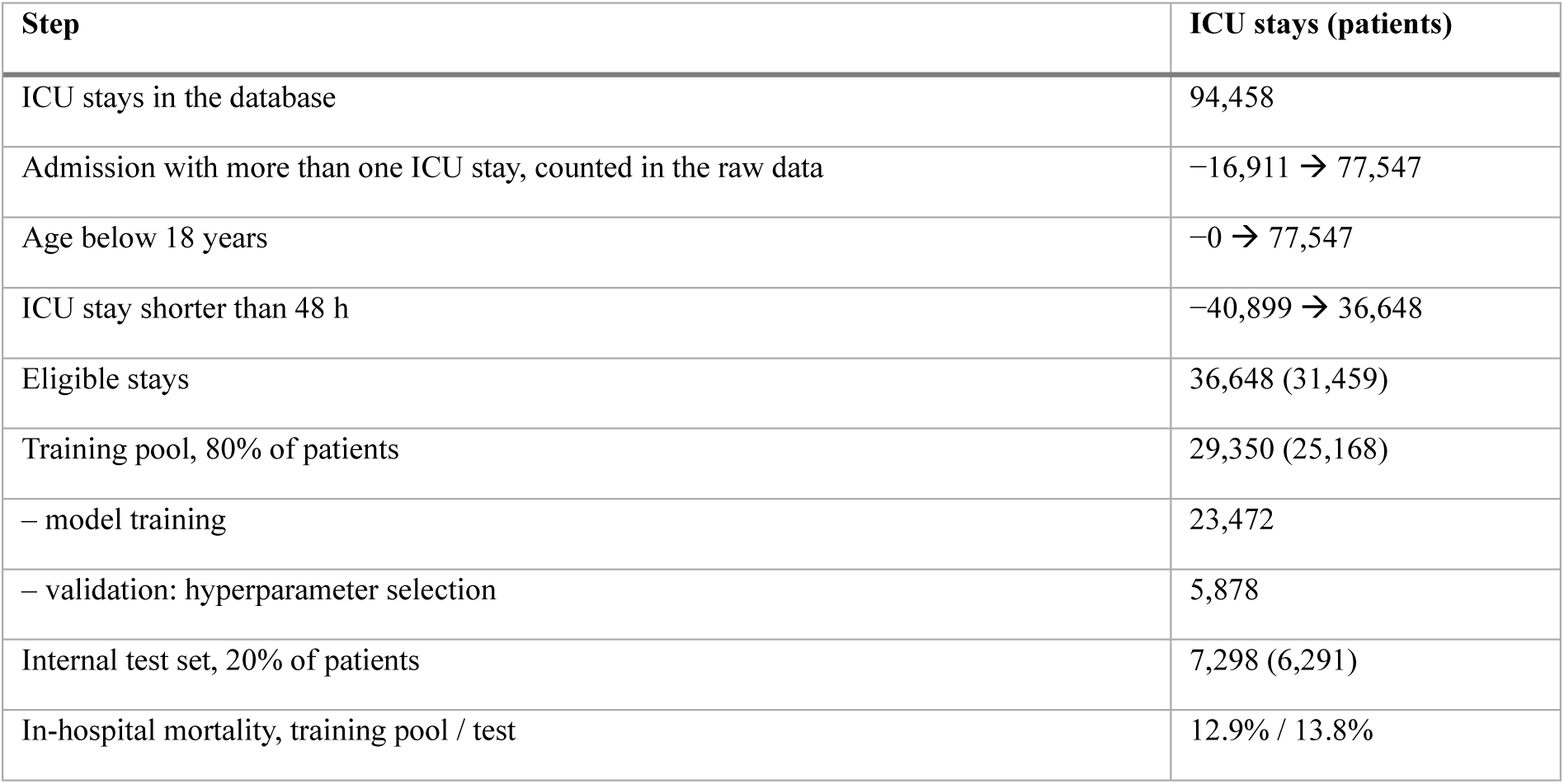
MIMIC-IV v3.1, development and internal test cohorts.

| **Step** | **ICU stays (patients)** |
|---|---|
| ICU stays in the database | 94,458 |
| Admission with more than one ICU stay, counted in the raw data | −16,911 → 77,547 |
| Age below 18 years | −0 → 77,547 |
| ICU stay shorter than 48 h | −40,899 → 36,648 |
| Eligible stays | 36,648 (31,459) |
| Training pool, 80% of patients | 29,350 (25,168) |
| – model training | 23,472 |
| – validation: hyperparameter selection | 5,878 |
| Internal test set, 20% of patients | 7,298 (6,291) |
| In-hospital mortality, training pool / test | 12.9% / 13.8% |

**Table 2.** eICU-CRD v2.0, external transfer cohort.

| **Step** | **ICU stays** |
|---|---|
| ICU stays in the database | 200,859 |
| Without a recorded hospital discharge status | −1,751 → 199,108 |
| Age below 18 years | −618 →198,490 |
| ICU stay shorter than 48 h | −121,537 → 76,953 |
| Admission with more than one ICU stay, counted after the filters above | −10,019 → 66,934 |
| External transfer cohort | 66,934 |
| In-hospital mortality | 10.9% |

No training or validation; all included stays were used for evaluation only.

### Study population

Of 94,458 ICU stays in MIMIC-IV, we excluded 16,911 stays from admissions with more than one ICU stay and 40,899 stays shorter than 48 hours, leaving 36,648 stays from 31,459 patients; MIMIC-IV contains adults only. The training pool comprised 29,350 stays from 25,168 patients and the internal test set 7,298 stays from 6,291 patients, with in-hospital mortality of 12.9% and 13.8% (Table 1).

In eICU-CRD, 1,751 of 200,859 ICU stays were first excluded because no hospital discharge status was recorded, so that no outcome can be derived. A further 618 stays were excluded for age below 18 years, 121,537 for an ICU stay shorter than 48 hours, and 10,019 for belonging to an admission with more than one ICU stay, leaving 66,934 stays with in-hospital mortality of 10.9% (Table 2).

The single-stay criterion was applied at a different point in the two databases. In MIMIC-IV, ICU stays per admission were counted in the raw data, so that an admission with one short and one long ICU stay was excluded entirely; in eICU-CRD, they were counted after the other criteria, so that the long stay of such an admission was retained. As a result, 10,378 external stays (15.5%) belong to admissions that the MIMIC-IV rule would have excluded. Restricting the external cohort to the remaining 56,556 stays changed no finding (Supplementary Table 2).

### Model performance after transfer

Discrimination declined in all models after transfer. On the internal test set, the AUROC was between 0.858 and 0.873; after transfer it fell to 0.762–0.773, a loss close to 0.10 in every architecture, with seed intervals that excluded zero (Table 3). On the precision–recall scale, the AUPRC fell from 3.7 to 4.2 times the mortality rate to 3.1 to 3.5 times. Deep ensembles of the ten seed models raised the AUROC by 0.004 to 0.009 but lost as much after transfer as single models.

**Table 3.** Mortality discrimination and calibration in-domain and after external transfer, three model architectures and their deep ensembles.

| Model | AUROC, in-domain | AUROC, transfer | Paired loss [seed interval] | AUPRC lift, in-domain / transfer | Slope, in-domain | Slope, transfer |
|---|---|---|---|---|---|---|
| GRU | 0.873 | 0.773 | 0.100 [0.098; 0.102] | 4.15 / 3.48 | 0.96 | 0.64 |
| LSTM | 0.868 | 0.762 | 0.105 [0.102; 0.109] | 4.10 / 3.38 | 0.92 | 0.57 |
| Transformer | 0.858 | 0.766 | 0.092 [0.090; 0.094] | 3.73 / 3.14 | 0.84 | 0.59 |
| Deep ensemble of the GRU | 0.877 | 0.777 | 0.100 | 4.22 / 3.54 | 1.01 | 0.67 |
| Deep ensemble of the LSTM | 0.872 | 0.767 | 0.105 | 4.17 / 3.44 | 0.96 | 0.60 |
| Deep ensemble of the transformer | 0.867 | 0.772 | 0.095 | 3.90 / 3.27 | 0.94 | 0.64 |

Means over ten seeds; brackets: 95% t-interval of the per-seed difference. Lift: AUPRC divided by cohort mortality. Slope: logistic recalibration on the predicted logit (ideal 1). Ensembles average the ten seed models’ probabilities. Calibration error for all seven configurations: Supplementary Table 5; cross-entropy: Supplementary Fig. 7.

The predicted risks also became too extreme after transfer: patients given a high risk died less often than predicted, and patients given a low risk died more often. This is measured by the calibration slope, which is 1 for a well-calibrated model and below 1 when predicted risks are more extreme than observed risks; it fell from 0.84 to 0.96 in-domain to 0.57 to 0.64 after transfer (Table 3, Fig. 1). Deep ensembles were better calibrated in-domain, with slopes of 0.94 to 1.01, but fell to 0.60 to 0.67 after transfer. The decline has the same origin as the loss of discrimination: the spread of the risk estimates within the outcome groups was unchanged, but survivors and non-survivors moved closer together, so that estimates of the same size were too extreme (Supplementary Note 5). On average, the models underestimated mortality to a similar degree in both cohorts. For every 100 deaths the GRU expected, 133 occurred in MIMIC-IV and 134 in eICU-CRD; for the LSTM, 119 and 119; for the transformer, 113 and 112. The transfer therefore changed how extreme the individual risk estimates were, not their overall level.

**Fig 1.**
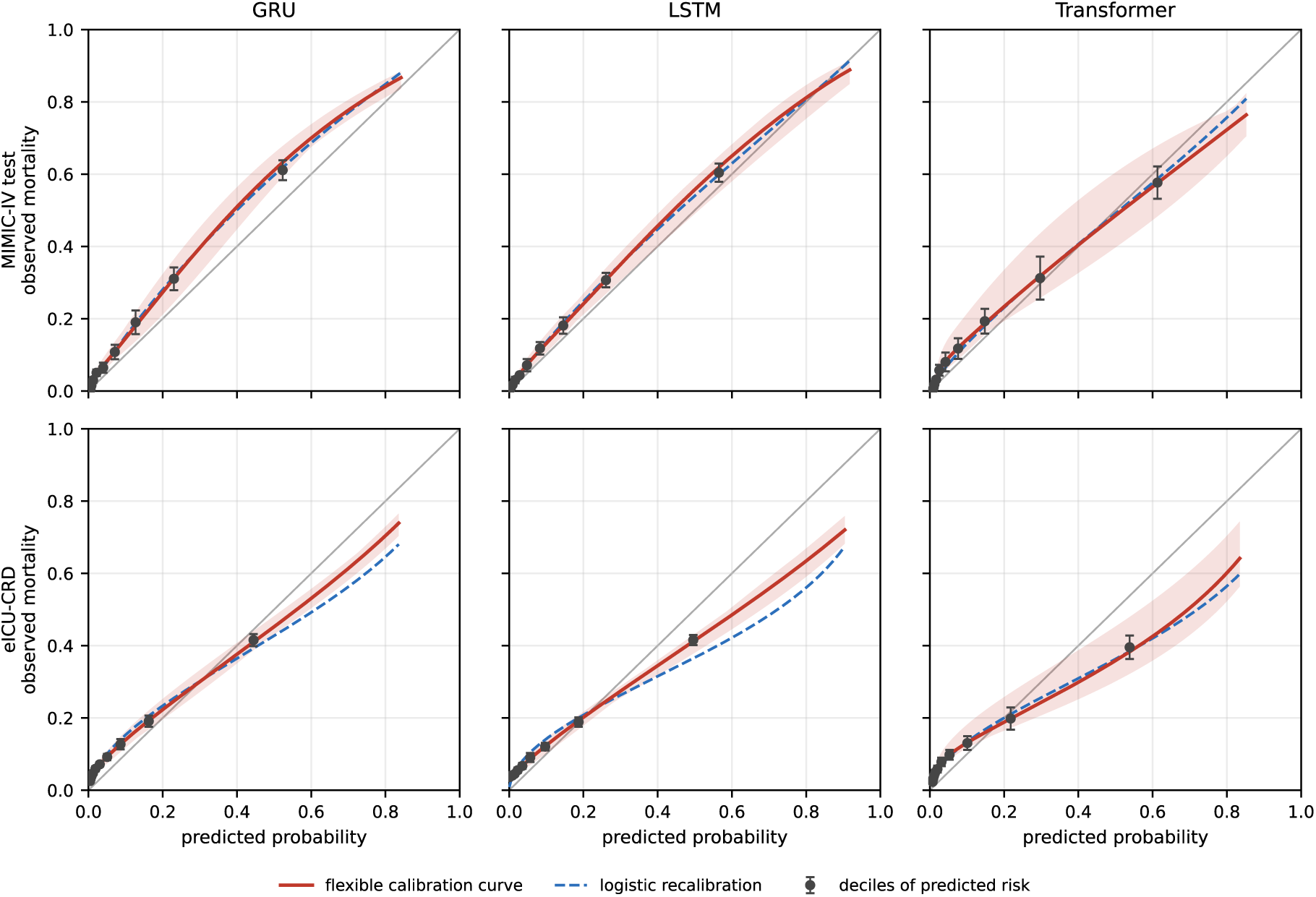
Calibration before and after transfer. Observed mortality against predicted probability for the three architectures on the internal test set (top) and the external cohort (bottom): the logistic recalibration line, and a flexible spline curve with its range over seeds. Decile points show means over seeds with ±1 SD. Curves stop below 1.0 because almost no patients received higher predicted probabilities; decile points end lower because each decile contains one tenth of the patients.

### Uncertainty after transfer

Predictive entropy and mutual information did not rise after transfer; both were slightly lower for patients in the external hospital system, and only representation density and the logit variance were weakly higher. For each uncertainty measure, we asked how often a randomly chosen external patient had a higher value than a randomly chosen internal patient, with 0.5 corresponding to chance (Supplementary Table 6).

The predictive entropy of the single models separated the cohorts at 0.475, 0.466, and 0.462 for the GRU, LSTM, and transformer, below chance in every architecture and every seed. The mutual information, which measures how much the repeated predictions for the same patient vary, was also lower on the external cohort: 0.477, 0.476, and 0.464 for Monte Carlo dropout and 0.474, 0.488, and 0.457 for deep ensembles, respectively. The total predictive uncertainty of both procedures separated the cohorts similarly weakly and in the same direction. Mutual information accounted for 0.9% to 2.7% of the total predictive uncertainty in both cohorts (mean mutual information divided by mean total uncertainty).

Only two uncertainty measures were higher on the external cohort, and only weakly: the representation density, at 0.516, 0.531, and 0.512, with seed intervals above 0.5, and the logit-variance component of Monte Carlo dropout, at 0.527, 0.533, and 0.533 (Fig. 2, Supplementary Table 6, Supplementary Fig. 2).

**Fig 2.**
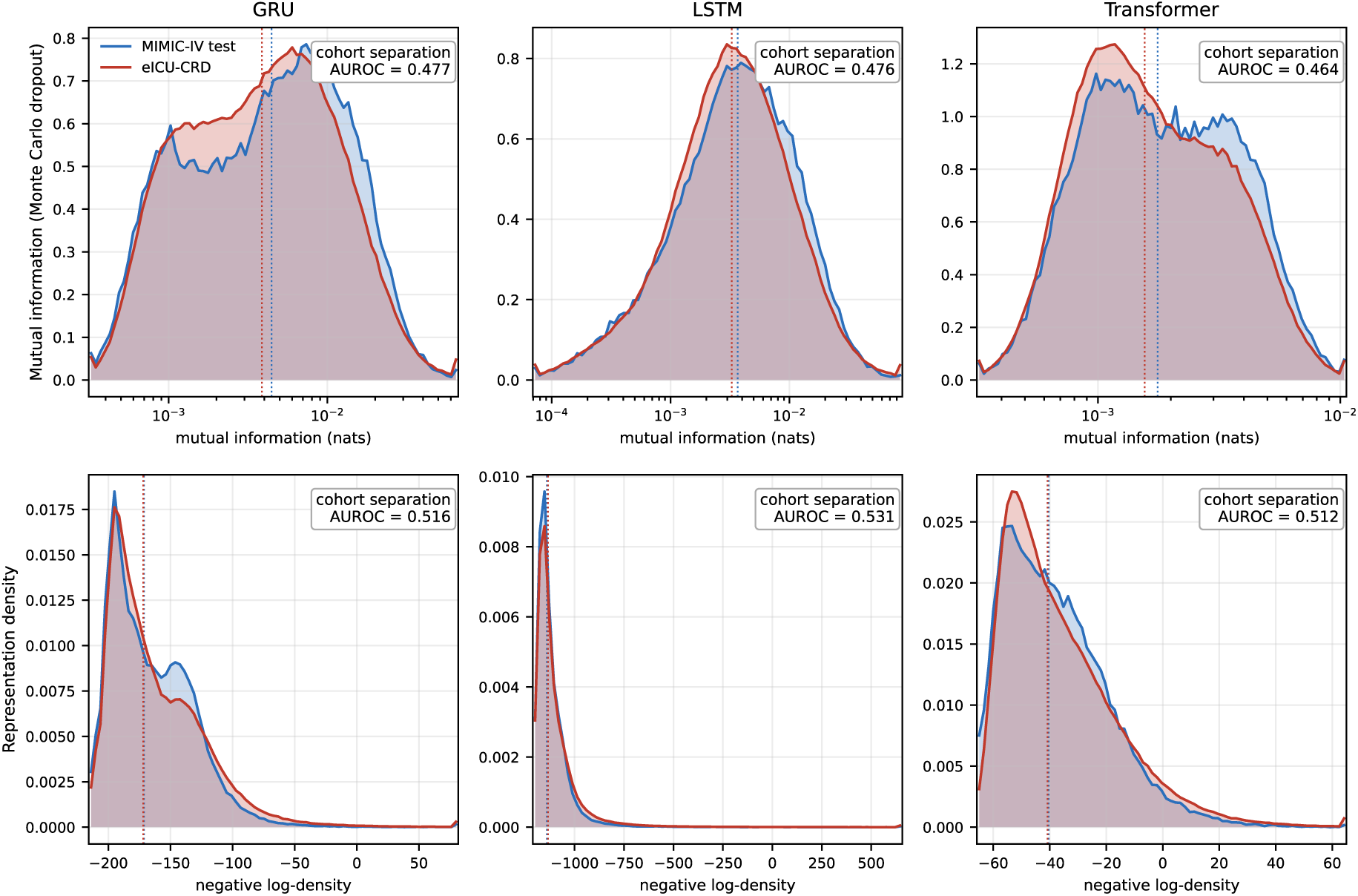
Mutual information and representation density. Top row: mutual information from Monte Carlo dropout, a measure of epistemic uncertainty, on a logarithmic x-axis. Bottom row: negative log-density, a measure of distance in the representation space of the last layer (higher values indicate greater distance from the centers of the training data). Blue: internal test set; red: external cohort. Dotted vertical lines: median of each cohort, with the ten seed models pooled. Boxes: cohort separation (AUROC, mean over the ten seed models).

### Uncertainty-based identification of prediction errors

Within the external cohort, the patients with the highest predictive entropy were the most likely to be misclassified. The entropy of the single models identified errors with AUROCs of 0.856, 0.858, and 0.870 for the GRU, LSTM, and transformer. Representation density and the logit-variance component, the only two uncertainty measures that had been slightly higher overall in the new hospital system, were less useful for spotting individual errors: the density identified errors less well than the entropy, and the logit variance tended to be higher for correct than for wrong predictions (Supplementary Table 6). The predictive entropy of Monte Carlo dropout reached 0.857, 0.861, and 0.869, within 0.003 of the single model in every architecture.

The epistemic components did less well. Mutual information reached 0.810, 0.791, and 0.836 for Monte Carlo dropout and 0.780 to 0.855 for deep ensembles. The representation density reached 0.789, 0.728, and 0.753; its paired difference to the predictive entropy ranged from −0.067 to −0.130 across all seven configurations and excluded zero in each (Supplementary Fig. 3). The logit-variance component showed inverse behavior, at 0.181 to 0.326: patients whose predictions varied most between passes were more often correctly classified.

### Deferring uncertain predictions

Deferring the most uncertain patients to clinical review left markedly fewer misclassifications among the remaining patients. With the 20% most uncertain patients deferred by the predictive entropy of the single model, the misclassification rate among the remaining patients fell from 17.5%, 18.9%, and 20.1% to 6.3%, 6.7%, and 8.0% for the GRU, LSTM, and transformer; with the predictive entropy of the deep ensemble, it fell to 6.0%, 6.1%, and 8.0% (Fig. 3; Supplementary Fig. 6).

**Fig 3.**
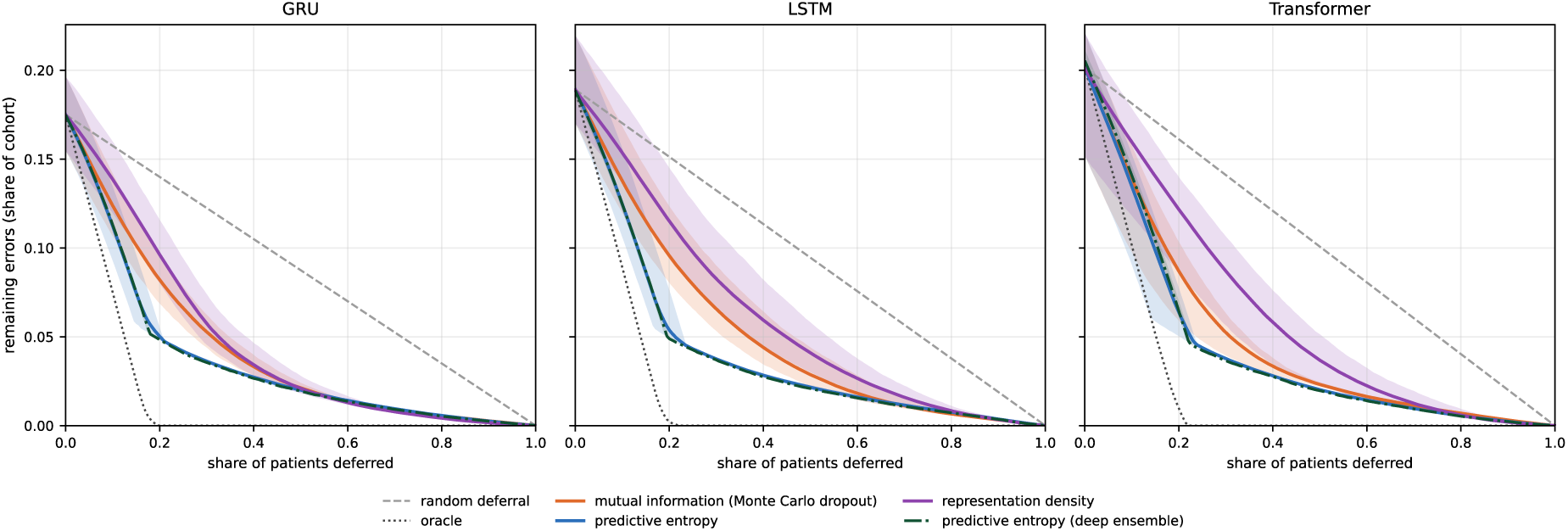
Deferring patients per architecture. The y-axis gives the errors remaining among the accepted patients as a share of the full cohort. Lines are means over ten seeds, shaded bands the range. Excess areas of all measures are given in Supplementary Table 6; the corresponding risk–coverage curves are shown in Supplementary Fig. 6.

The deferral captured 91% to 99% of false-positive predictions but only 2% to 7% of missed deaths, so that most misclassifications remaining after deferral were deaths at low predicted risk (Supplementary Table 9; Supplementary Fig. 8).

Across all shares of deferred patients, the predictive entropy of the single model left an excess area under the risk–coverage curve of 0.037 to 0.042 (Fig. 4; Supplementary Table 6). The deep ensemble improved on this by 0.0004 to 0.0012. The predictive entropy of Monte Carlo dropout left 0.037, 0.040, and 0.038, within 0.002 of the single model in every architecture. Deferring by mutual information (0.047 to 0.056), by representation density (0.048 to 0.072, 1.3 to 1.8 times the excess area of the entropy; Supplementary Fig. 4), or by the logit-variance component (0.273 to 0.365) was worse in every architecture.

**Fig 4.**
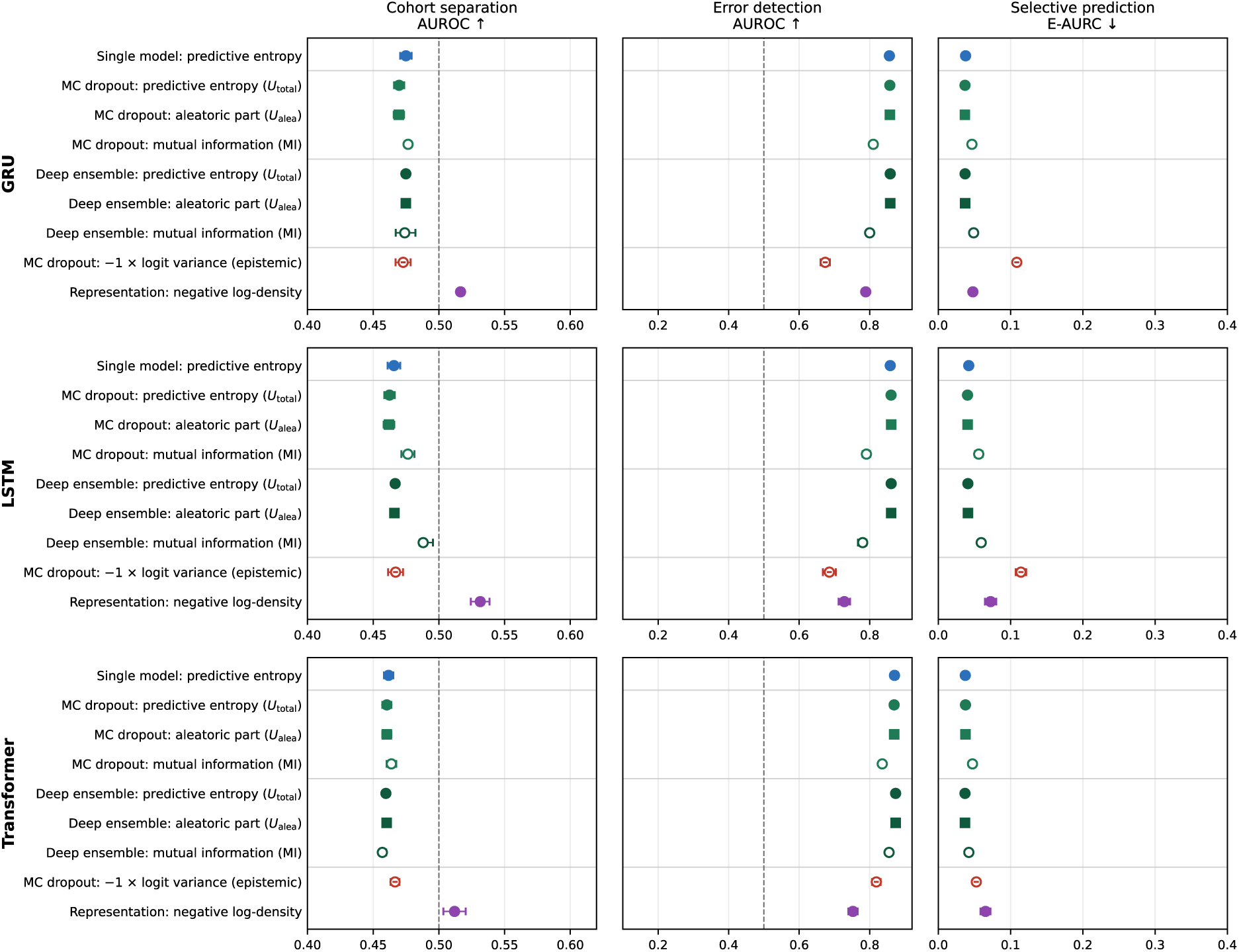
Uncertainty measures across cohort separation, error detection, and selective prediction. Patient-level analyses use the external cohort. Higher AUROC and lower excess area under the risk–coverage curve (E-AURC) indicate better performance. Points show means across ten training seeds with 95% t-intervals; ensembles show full-ensemble estimates with leave-one-member-out ranges. Errors are those of the deterministic model or, for ensemble measures, the ensemble mean (classification threshold τ = 0.1288, the mortality rate of the training pool). Dashed lines mark chance (AUROC 0.5). The logit variance is plotted with reversed sign (−1 × logit variance): its AUROCs here equal 1 minus the values in the Results and Supplementary Table 6, and its E-AURC corresponds to deferring the patients with the lowest logit variance first. MC, Monte Carlo.

On the internal test set the predictive entropy of the single model identified errors with AUROCs of 0.873, 0.878, and 0.878, that is 0.017, 0.020, and 0.007 higher than on the external cohort, and left an excess area under the risk-coverage curve of 0.026 to 0.028 against 0.037 to 0.042 after transfer (Supplementary Table 12). The ranking of the measures was the same in both cohorts.

Taken together, higher uncertainty after transfer did not imply better performance in error detection or selective prediction. Predictive entropy was lower in the external cohort but remained as the most useful measure for identifying errors and prioritizing predictions for review (Fig. 4).

### Sensitivity analyses

The constrained variants of the GRU and the LSTM shifted individual values, lowering the cohort separation of the entropy by 0.03 to 0.05 and raising that of the density by 0.02 overall, but changed no ordering: the entropy remained below chance, the density at most weakly above it, and at the patient level the density stayed below the entropy in every variant (Supplementary Table 6). Among the further density definitions, a shared covariance, a per-class Gaussian mixture, and the class-conditional Gaussian were above 0.5 in all seven configurations and the relative Mahalanobis distance of Ren et al.^26^ in five of seven, all below 0.57; a background-corrected relative density score was above 0.5 in two and a density on length-normalized representations in two (Supplementary Table 6, Supplementary Fig. 2).

Restricting the external cohort to the 56,556 stays that satisfy the MIMIC-IV admission criterion changed every uncertainty measure by less than 0.01 (Supplementary Table 2), and refitting the density on a class-balanced training subset changed its cohort separation by less than 0.01 (Supplementary Note 7). At a classification threshold of 0.5, every uncertainty measure identified errors less well. The Monte Carlo entropy and the single-model entropy stayed within 0.01 of each other at both thresholds (0.761 vs 0.755, 0.753 vs 0.744, and 0.770 vs 0.767 at 0.5), and the density on length-normalized representations exceeded the single-model entropy for the GRU (0.764 vs 0.755; Supplementary Table 8, Supplementary Note 2).

Finally, 66 of the 70 trained models had reached their lowest validation loss before the final epoch of the fixed training length, and assessing each base model at that epoch instead of the end of training changed no conclusion (Supplementary Note 10; Supplementary Table 10; Supplementary Table 11).

## Discussion

After transfer to another hospital system, discrimination and calibration declined, but no uncertainty measure rose to signal this loss: predictive entropy and mutual information were in fact lower in the external cohort. Within the external cohort uncertainty however identified the single predictions most likely to be wrong, and deferring the most uncertain patients removed most misclassifications, mostly false alarms, from the predictions that remained. The uncertainty measure that served best on both patient-level questions was the simplest one, the predictive entropy of the model itself; sampling-based and distance-based uncertainty measures added little or did worse. Uncertainty, as measured by predictive entropy, thus ranked predictions by their reliability but did not reveal that the model as a whole had become less reliable.

Most external patients received a low predicted risk of death (Supplementary Fig. 1); predictions near 0% or 100% have low entropy, whereas predictions near 50% have high entropy because survival and death are considered equally likely. Low entropy therefore indicates a more certain prediction, not a correct one: it decreased even though the models became less able to distinguish survivors from non-survivors and produced overly extreme risk estimates (calibration slopes below 1). Ovadia et al.^7^ showed that good calibration on the original data does not guarantee reliable uncertainty estimates when the data change; transfer between hospital systems is a clinical instance of this. Monitoring for an increase in entropy would have failed to flag the observed deterioration.

Disagreement between repeated predictions did not indicate the deterioration either; two properties of mutual information may explain why. It accounted for only a few percent of the total predictive uncertainty, and for the GRU and the transformer the difference between cohorts largely disappeared when patients were compared within deciles of predicted risk (Supplementary Note 9), so that it partly reflected the distribution of predicted risks. In addition, ensemble members and dropout passes may have made similar errors after transfer; agreement among repeated predictions does not imply that they are correct.

These results follow from what the measures estimate: entropy and mutual information describe the predicted outcome and can therefore rank errors but need not rise when the inputs change, whereas representation density describes the inputs and can register a change in the population but says nothing about whether a given prediction is correct. That the density separated the cohorts only weakly leaves two explanations: the two cohorts look alike in the learned representation, or the representation or the density estimator overlooked the changes that mattered for prediction. The outcome is in line with Postels et al.^27^, who found that strong detection of unfamiliar inputs by deterministic methods does not guarantee reliable uncertainty under continuous distribution shift, and contrasts with the image-benchmark results of Mukhoti et al.^28^, where such densities matched deep ensembles.

Predictive entropy still helped identify patient-level errors within the external cohort. Most correctly classified patients had very low predicted risks and low entropy. Misclassified patients often had intermediate predicted risks, where entropy is higher. When a fixed proportion of predictions is selected for review, what matters is whether incorrect predictions rank above correct ones. Entropy can therefore help select individual predictions for review even when its overall level is lower in the external cohort. Compared with the internal test set, error detection by entropy lost 0.007 to 0.020 in AUROC and the excess area rose by about a third. After transfer, entropy still separated incorrect from correct predictions almost as well as at the development site, but it gave no indication that the model had moved to a hospital system where it performed worse.

Our main Gaussian density score identified errors best in the GRU, where it ranked patients most similarly to entropy. The density based on length-normalized representations also followed entropy closely and performed best among the density measures (Supplementary Table 7).

High logit variance, which measures variation in the model’s internal scores across dropout passes, tended to prioritize correct predictions for review (Supplementary Note 9). High values often occurred at very low or very high predicted risks, where most classifications were correct. At these extremes, substantial differences in internal scores can translate into only small differences in predicted probabilities. This may partly explain why logit variance was a poor indicator of prediction errors.

The most actionable finding is that predictive entropy ranked errors within the external cohort and supported selective deferral: it requires no additional training or repeated passes, can be reported with every prediction^13^, and reviewing one in five patients would have reached about 70% of all misclassifications (Fig. 5; Supplementary Fig. 8). A prespecified entropy threshold, informed by the entropy on the internal test cohort, could trigger review of the patient record, missing measurements, and model output before the prediction informs care. Whether entropy ranks errors in a new setting, and at which threshold, can itself only be verified once outcomes are known, as for calibration; in our transfer this property carried over with modest loss (Supplementary Table 12), but that is not guaranteed elsewhere.

**Fig 5.**
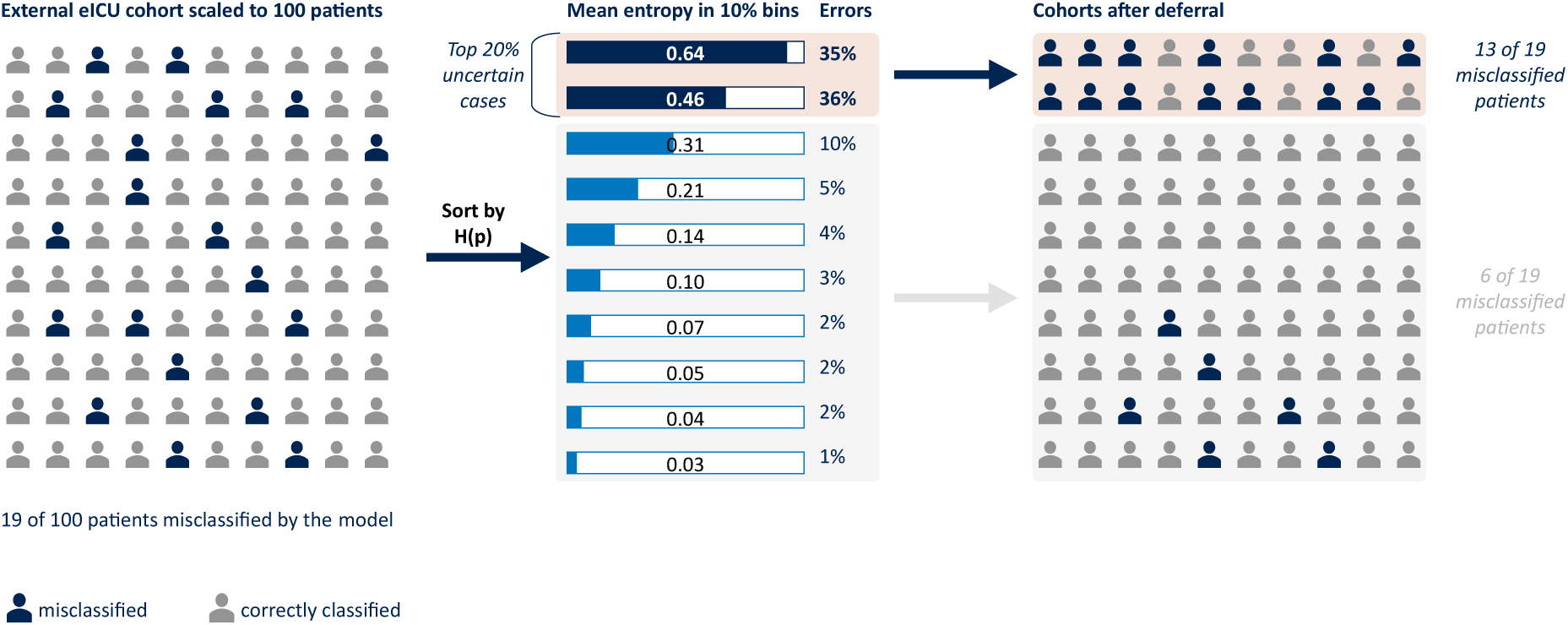
Deferral per 100 patients. Patients sorted by predictive entropy, H(p), into bins of ten (single-model entropy, means over the three architectures and ten seeds). Errors: share of all misclassifications falling in the bin. The 19 misclassified patients per 100 are the mean misclassification rate of the three architectures (GRU 17.5%, LSTM 18.9%, transformer 20.1%).

Such a workflow could contribute to human oversight of high-risk AI systems under Article 14 of the EU AI Act^16^. The threshold should be chosen against an acceptable residual error rate and available review capacity and the rule must state which errors it targets: because entropy is highest at a predicted risk of 50% and low near 0%, the rule evaluated here selected almost exclusively false alarms, while most remaining errors were deaths among patients classified as low risk (Supplementary Table 9; Supplementary Fig. 8). Such unflagged errors could create false reassurance, and low entropy should not waive critical review, especially when other clinical concerns are present. How such information is presented matters: limited AI literacy has been identified as a barrier to using model transparency information for oversight^29^, which makes AI competencies an essential part of medical training^30^.

Post-market monitoring should not rely solely on the model’s own uncertainty. Article 72 of the EU AI Act and Article 83 of the Medical Device Regulation require ongoing monitoring of high-risk AI systems and medical devices, respectively^16,17^. Finlayson et al.^31^ described how changes in patient populations, clinical practice, and data recording can undermine clinical AI systems. Our findings add that poorer performance need not be accompanied by increases in entropy or mutual information, but predictive entropy is a valuable uncertainty measure before true outcomes are known.

Monitoring should include regular checks of patient characteristics, missing measurements, and data recording practices, which can begin before outcomes are known. Once outcomes become available, planned audits should assess how well the model still separates patients with and without the outcome of the event that the model predicts, how closely predicted risks match observed outcomes across the risk range, and how often clinically important errors occur. These audits should specifically examine deaths among patients classified as low risk and check whether the review rule still selects the errors it was designed to identify. Shared terminologies and standards-based repositories such as openEHR support data interoperability across institutions^32^. However, harmonized data structures do not ensure model generalizability, making local validation and ongoing monitoring essential.

Although representation density separated the cohorts only weakly, tracking its distribution across many patients might help identify changes that warrant investigation. Whether this provides a timely and reliable monitoring measure, including for changes within individual hospitals^33^, remains to be tested. As the models themselves gave no such clue, information on the model’s development population may help clinicians assess whether it is being applied to patients and clinical settings similar to those in which it was developed^34^.

This study has three main limitations. First, its scope: it examined a single prediction task, in-hospital mortality from the first 48 hours, a single transfer between two US critical care databases, and 13 routinely charted variables, with glucose and pH as the only laboratory values and no medications. The evaluation framework applies to other settings, but whether the findings, including the deferral results, hold for other outcomes, health systems, and richer inputs remains to be tested.

Second, the comparison of uncertainty methods is bounded in two ways. Other distance-aware approaches, such as spectral-normalized Gaussian processes^35^ and deterministic uncertainty quantification^36^, were not evaluated, and the encoder constraints bound distances only across the final layer, not end to end.

Third, the design is retrospective and the statistics describe model variability, not patient sampling. Intervals reflect variability across training runs on fixed cohorts; differences between the hospitals within eICU-CRD were not analyzed, and the breakdown of deferred errors was conducted after the main results were known. Selective prediction was evaluated by deferring the most uncertain patients and measuring errors among the remaining ones; the consequences of false alarms and missed deaths were not weighed against each other, and how such deferral would affect clinical decisions in prospective use was not assessed.

After transfer between hospital systems, model discrimination and calibration worsened while predictive entropy decreased. Representation density only weakly distinguished patients from the two cohorts, whereas predictive entropy was more useful for identifying errors and selecting predictions for review within the external cohort. These findings highlight the distinction between detecting changes in the patient population and identifying individual prediction errors. A model whose performance has worsened may nevertheless appear more confident.

Predictive entropy could help prioritize clinical review, but lower error rates among predictions not selected for review do not establish that review improves clinical decisions. Under the review rule evaluated here, most remaining errors involved patients who died despite being classified as low risk. Clinical use therefore requires validation in the receiving hospital and prospective evaluation of review thresholds and workflows, including which errors remain undetected and how much clinician time review requires. Model performance should continue to be monitored after deployment as patient outcomes become available.

## Methods

### Study design and data sources

In this retrospective study we developed in-hospital mortality prediction models with MIMIC-IV v3.1 as the development database and eICU-CRD v2.0 as an external transfer cohort^37,38^. Both are de-identified critical care databases from hospitals in the United States and are available through PhysioNet under credentialed access. Both cohorts were restricted to adults, ICU stays of at least 48 hours, and hospital admissions with a single ICU stay; a patient with several hospital admissions contributes each of them. MIMIC-IV patients were split at random, with 20% held out as the internal test set so that no patient contributes to both sets; the remaining 80% formed the training pool, part of which served as the validation split for hyperparameter selection. All included eICU-CRD stays were used for evaluation only.

### Outcome and predictors

The outcome was in-hospital mortality, predicted from the first 48 hours of the ICU stay, following the benchmark task of Harutyunyan et al^39^. Thirteen routinely charted variables were extracted for each of the 48 hours (Supplementary Table 1). Multiple measurements within one hour were averaged, as in MIMIC-Extract^40^ rather than represented by the last measurement as in the logic of Harutyunyan et al. Missing hours were filled with the last available value of the same patient, hours before the first measurement with the first observed value, and variables never measured with a clinical default^41^. Continuous variables were standardized with the mean and standard deviation of the MIMIC-IV training data, and the same values were applied to eICU-CRD; standardizing the external cohort with its own statistics would have removed part of the shift the study is designed to measure. The in-hospital mortality rate of the training pool was 12.88% (Table 1).

### Model development

We compared three base model architectures that are widely used for clinical time series^42,43^; a long short-term memory network (LSTM), which is a baseline of mortality benchmarks^39,44^, a gated recurrent unit (GRU), the second widely used recurrent variant for irregularly sampled clinical time series^45^ and a transformer encoder^46^, which relates all hours of a stay to each other instead of reading them in sequence. Each architecture condenses the 48 hours into an internal representation of the patient, from which a final layer computes the predicted probability.

One of the uncertainty measures, the representation density, scores a patient by how close the patient’s internal representation lies to those of the training cohort. This assumes that patients who differ in their inputs also differ in the model’s representations^36^. To explore whether enforcing this property has an impact on the results, we trained additional variants of the GRU and LSTM in which spectral normalization limits how strongly the network can stretch differences between patients in the representation space, and a residual block limits how strongly it can compress them^28,35,47^. The residual block is the last layer before the prediction head and bounds distances only across this layer, not across the recurrent layers before it (Supplementary Note 3).

Models were trained with the Adam optimizer^48^ and binary cross-entropy. Hyperparameters, including the number of passes through the training data (epochs, 10 to 40), were selected per architecture with Optuna^49^ via a validation split of the training pool (Supplementary Table 3, Supplementary Table 4, Supplementary Note 4); the constrained variants used those of their base architecture. Each model was trained ten times with different random seeds, and the ten models of an architecture were also combined into a deep ensemble by averaging their predicted probabilities^50^. Through Monte Carlo dropout, we obtain a predictive distribution for each patient using 30 dropout masks^51^. The passes used the trained deterministic models themselves, with every dropout layer that was active in training also active at prediction time. The dropout uncertainty measures were calculated from the 30 predicted mortality probabilities and their corresponding log-odds. The equations for all components are given in Supplementary Note 1.

Each model returns an estimated probability that the patient dies in hospital. A prediction was classified as positive if this probability exceeded 0.1288, the mortality rate of the training pool. A prediction was therefore misclassified if the classification differed from the observed outcome. A threshold of 0.5 was examined as a sensitivity analysis in Supplementary Note 2.

### Uncertainty measures

We compared three uncertainty measures, all computed per patient.

The first is the predictive entropy of a single model. It is highest when the predicted probability is near 0.5 and zero when the model is certain:

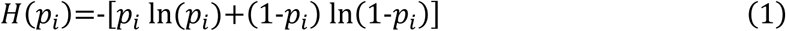

The second comes from repeating a prediction either through T=30 Monte Carlo dropout passes or through the T=10 members of a deep ensemble. With *p*^(*t*)^*_i_* being the prediction of pass or member *t*, the entropy of the averaged prediction is split into a part that the repeated predictions share and a part that reflects their disagreement^51,52^:

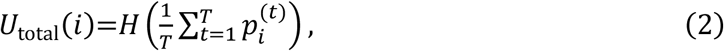

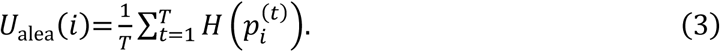

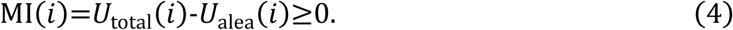

Total predictive uncertainty (*U*_total_) is the entropy of the averaged mortality prediction. The aleatoric component (*U*_alea_) is the average entropy of the individual predictions and represents uncertainty within those predictions. Their difference, mutual information (MI), measures disagreement between predictions and serves as the epistemic uncertainty measure. MI is zero when all predictions agree, even if each prediction remains uncertain. These definitions apply to both Monte Carlo dropout and deep ensembles. For Monte Carlo dropout, we also calculated the logit variance, the variance of the log-odds across the 30 passes, as a second measure of epistemic uncertainty (Supplementary Note 1, Eq. S7).

The third kind, representation density, is computed from a single forward pass of a single model. Following deep deterministic uncertainty^28^, a Gaussian model with one component per outcome class is fitted to the internal representations ℎ of the training patients. We used the negative logarithm of the estimated density as the uncertainty score, with higher values indicating greater unfamiliarity:

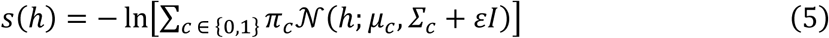

with class frequencies *π_c_*, means *μ_c_* and covariances *Σ_c_*; high values of *s*(ℎ) indicate a patient unlike the training data. Five further variants were evaluated on the same representations: a covariance shared across classes, a three-component mixture per class, the density on length-normalized representations^53^, a background-corrected relative density score, and the relative Mahalanobis distance of Ren et al.^26^ (Supplementary Note 7). The density was also refitted on a class-balanced subset of the training pool as a sensitivity analysis.

### Evaluation

Discrimination was measured by the area under the receiver operating characteristic curve (AUROC) and by the area under the precision–recall curve relative to the mortality rate (AUPRC lift). The primary calibration measure was the calibration slope, estimated by a logistic regression of each patient’s observed outcome on the model’s log-odds, without grouping patients into bins^6,54^. A slope of 1 is ideal; below 1, high predicted risks are too high and low predicted risks too low. A linear fit on the deciles of Fig. 1 served as a sensitivity analysis (Supplementary Note 5).

We also report the expected calibration error over deciles of the predicted risk (ECE; Supplementary Note 6)^55,56^ and the cross-entropy relative to a constant prediction of the mortality rate (skill; Supplementary Fig. 7).

Each uncertainty measure was evaluated at three levels, corresponding to each of the research questions:

Level 1, cohort separation, is the AUROC of the measure for distinguishing external from internal test patients: the probability that a randomly chosen external patient has the higher value, with 0.5 corresponding to chance and values above 0.5 to a measure that rises after transfer.

Level 2, error detection, is the AUROC of the measure for distinguishing misclassified from correctly classified external patients. Errors were those of the prediction used together with the uncertainty measure: for single-model and Monte Carlo measures those of the model’s own prediction without dropout, for ensemble measures those of the ensemble mean.

Level 3, selective prediction, asks whether deferring the most uncertain patients to clinical review reduces the errors among those who remain. Patients were accepted from the most certain downward, and the misclassification rate among the *k* accepted patients, *R*(*k*), was averaged over all *k* to give the area under the risk–coverage curve (AURC)^57,58^. Subtracting the area of an oracle that accepts all correctly classified patients first removes the effect of the overall error rate^59^:

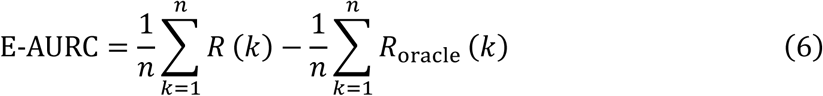

Lower values are better, and 0 is ideal. We also report the misclassification rate among the accepted patients when the 20% most uncertain are deferred (risk at 80% coverage). Patients with identical uncertainty measure values were accepted in the order of the cohort file, so that the result is reproducible (Supplementary Note 8).

Levels 2 and 3 were also computed on the internal test set as a baseline without distribution shift (Supplementary Table 12). Together, the three levels correspond to the two use cases examined in this study: Level 1 tests whether a measure warns of poorer overall performance after transfer, and Levels 2 and 3 whether it identifies the predictions that need review.

Table 4 summarizes what each measure quantifies and which question it addresses directly; here P(Y | X) denotes the probability of the outcome Y given a patient’s inputs X, which the model estimates, and P(X) the distribution of the inputs themselves.

**Table 4.** Uncertainty measures, what they quantify, and the research question each addresses directly.

| Measure | What it quantifies | Addresses directly |
|---|---|---|
| Predictive entropy (single model, Monte Carlo dropout, deep ensemble) | Uncertainty of the predicted outcome, P(Y \| X): highest near 50% risk, zero near 0% or 100% | Error detection and selective prediction<br>(questions 2 and 3) |
| Mutual information; logit variance | Disagreement between repeated predictions of<br>P(Y \| X), the epistemic part | Error detection and selective prediction (questions 2 and 3); cohort separation only if disagreement rises for unfamiliar inputs |
| Representation density | Familiarity of the input, P(X), estimated in the model’s representation space | Cohort separation<br>(question 1) |

### Use of large language models

A large language model was used during manuscript preparation solely for language editing, proofreading, and for writing analysis code to the authors’ specifications, which the authors reviewed. It was not used to generate scientific content, perform analyses, interpret results, or draft the manuscript independently. No patient-level data were shared with any AI service; the models operated on aggregated results only, in line with the PhysioNet data use agreement. The authors reviewed all AI-assisted edits and take full responsibility for the final content.

### Ethics

This study is a secondary analysis of two publicly available, de-identified critical care databases. MIMIC-IV was approved by the institutional review boards of the Massachusetts Institute of Technology and Beth Israel Deaconess Medical Center, and eICU-CRD was released under the HIPAA safe harbor provision after certification of its de-identification^37,38^. Both are available under a data use agreement.

## Data availability

The used databases, MIMIC-IV v3.1 and eICU-CRD v2.0, are available from PhysioNet (https://physionet.org/content/mimiciv/3.1/ and https://physionet.org/content/eicu-crd/2.0/) to credentialed users under a data use agreement. The derived cohorts cannot be shared under the agreement but can be regenerated from the raw data with the code below. The aggregated results underlying all figures and tables are provided in the Supplementary Information and in the code repository.

## Code availability

The underlying code for this study is available in the GitHub repository https://github.com/stebehling/uncertainty-icu-transfer.

## Acknowledgements

We would like to acknowledge the preparatory work of Lukas Scholz during his M.Sc. thesis.

This work received no specific funding from any agency in the public, commercial, or not-for-profit sectors.

## Author contributions

SB: Conceptualization, data curation, formal analysis, software, investigation, methodology, project administration, visualization, writing—original draft and revision.

MSA: Writing—review and editing.

SH: Methodological guidance, review and editing. EG: Review and editing.

FB: Clinical guidance, review and editing.

NF: Conceptualization, data curation, formal analysis, investigation, methodology, project administration, visualization, writing—original draft and revision.

SDB: Conceptualization, data curation, formal analysis, investigation, methodology, project administration, visualization, writing—original draft and revision.

## Competing interests

The authors declare no competing financial or non-financial interests.

## Notes

### Competing Interest Statement

The authors have declared no competing interest.

### Author Declarations

The Institutional Review Boards of Beth Israel Deaconess Medical Center (Boston, MA, USA) and the Massachusetts Institute of Technology (Cambridge, MA, USA) gave ethical approval for the collection and de-identification of the MIMIC-IV database and waived the requirement for individual informed consent. The eICU Collaborative Research Database was released by Philips Healthcare and the MIT Laboratory for Computational Physiology as a de-identified database exempt from institutional review board approval; its de-identification was certified under the HIPAA Safe Harbor provision (Privacert, Cambridge, MA, USA). Both databases were accessed under the PhysioNet credentialed health data use agreement after completion of the CITI "Data or Specimens Only Research" course. This study is a secondary analysis of these de-identified data without patient contact or additional data collection and required no additional ethics approval.

## References

1. Rajkomar A, Oren E, Chen K, et al. Scalable and accurate deep learning with electronic health records. Npj Digit Med. 2018;1(1):18. doi:10.1038/s41746-018-0029-1

2. Adams R, Henry KE, Sridharan A, et al. Prospective, multi-site study of patient outcomes after implementation of the TREWS machine learning-based early warning system for sepsis. Nat Med. 2022;28(7):1455–1460. doi:10.1038/s41591-022-01894-0

3. Boie SD, Giesa N, Sekutowicz M, et al. Multimodal data for predictive medicine: algorithmic fusion of clinical data in anesthesiology and intensive care. Front Med. 2026;13:1746867. doi:10.3389/fmed.2026.1746867

4. Rockenschaub P, Hilbert A, Kossen T, et al. The Impact of Multi-Institution Datasets on the Generalizability of Machine Learning Prediction Models in the ICU. Crit Care Med. 2024;52(11):1710-1721. doi:10.1097/CCM.0000000000006359

5. Rockenschaub P, Akay EM, Carlisle BG, et al. External validation of AI-based scoring systems in the ICU: a systematic review and meta-analysis. BMC Med Inform Decis Mak. 2025;25(1):5. doi:10.1186/s12911-024-02830-7

6. Van Calster B, McLernon DJ, Van Smeden M, Wynants L, Steyerberg EW. Calibration: the Achilles heel of predictive analytics. BMC Med. 2019;17(1):230. doi:10.1186/s12916-019-1466-7

7. Ovadia Y, Fertig E, Ren J, et al. Can You Trust Your Model’s Uncertainty? Evaluating Predictive Uncertainty Under Dataset Shift. *arXiv*. Preprint posted online December 17, 2019:arXiv:1906.02530. doi:10.48550/arXiv.1906.02530

8. Boie SD, Reis F, Frey N, Grünewald E, Balzer F. Calibration of Self-Reported Confidence and Accuracy of Large Language Models in Medical Question Answering. J Med Syst. 2026;50(1):103. doi:10.1007/s10916-026-02430-0

9. Wong A, Otles E, Donnelly JP, et al. External Validation of a Widely Implemented Proprietary Sepsis Prediction Model in Hospitalized Patients. JAMA Intern Med. 2021;181(8):1065–1070. doi:10.1001/jamainternmed.2021.2626

10. Antoniadi AM, Du Y, Guendouz Y, et al. Current Challenges and Future Opportunities for XAI in Machine Learning-Based Clinical Decision Support Systems: A Systematic Review. Appl Sci. 2021;11(11):5088. doi:10.3390/app11115088

11. Frey N, Agha-Mir-Salim L, Renner L, Haufe S, Voge L, Balzer F. Clinical explainable AI evaluations prioritize trust over effective human oversight: a scoping review. Research Square. Preprint posted online September 24, 2026. doi:10.21203/rs.3.rs-10892043/v1

12. Ghassemi M, Oakden-Rayner L, Beam AL. The false hope of current approaches to explainable artificial intelligence in health care. Lancet Digit Health. 2021;3(11):e745–e750. doi:10.1016/S2589-7500(21)00208-9

13. Frey N, Giesa N, Agha-Mir-Salim L, Balzer F. Physicians’ Perspectives on Predictive Uncertainty in Machine Learning Models. In: Giacomini M, Delgado J, Arvanitis TN, et al., eds. Studies in Health Technology and Informatics. IOS Press; 2026. doi:10.3233/SHTI260277

14. Kompa B, Snoek J, Beam AL. Second opinion needed: communicating uncertainty in medical machine learning. Npj Digit Med. 2021;4(1):4. doi:10.1038/s41746-020-00367-3

15. Frey N, Meyer-Eschenbach F, Kumpf O, et al. Predictive uncertainty representations in clinical AI shape physicians’ advice uptake, accuracy, and trust. Commun Med. 2026;in press.

16. Regulation (EU) 2024/1689 of the European Parliament and of the Council of 13 June 2024 laying down harmonised rules on artificial intelligence and amending Regulations (EC) No 300/2008, (EU) No 167/2013, (EU) No 168/2013, (EU) 2018/858, (EU) 2018/1139 and (EU) 2019/2144 and Directives 2014/90/EU, (EU) 2016/797 and (EU) 2020/1828 (Artificial Intelligence Act). OJ L, 2024/1689, 12.7.2024.

17. Regulation (EU) 2017/745 of the European Parliament and of the Council of 5 April 2017 on medical devices, amending Directive 2001/83/EC, Regulation (EC) No 178/2002 and Regulation (EC) No 1223/2009 and repealing Council Directives 90/385/EEC and 93/42/EEC. OJ L 117, 5.5.2017, p. 1–175.

18. Siontis GCM, Tzoulaki I, Castaldi PJ, Ioannidis JPA. External validation of new risk prediction models is infrequent and reveals worse prognostic discrimination. J Clin Epidemiol. 2015;68(1):25–34. doi:10.1016/j.jclinepi.2014.09.007

19. Harrison DA, Brady AR, Parry GJ, Carpenter JR, Rowan K. Recalibration of risk prediction models in a large multicenter cohort of admissions to adult, general critical care units in the United Kingdom. Crit Care Med. 2006;34(5):1378–1388. doi:10.1097/01.CCM.0000216702.94014.75

20. Azizmalayeri M, Abu-Hanna A, Cinà G. Unmasking the chameleons: A benchmark for out-of-distribution detection in medical tabular data. Int J Med Inf. 2025;195:105762. doi:10.1016/j.ijmedinf.2024.105762

21. Bernhardt M, Ribeiro FDS, Glocker B. Failure Detection in Medical Image Classification: A Reality Check and Benchmarking Testbed. Trans Mach Learn Res. Published online 2022. https://openreview.net/forum?id=VBHuLfnOMf

22. Lopez LJL, Shamout FE, Rudner TGJ. An Empirical Analysis of Calibration and Selective Prediction in Multimodal Clinical Condition Classification. In: Proceedings of the 7th Conference on Health, Inference, and Learning. Vol 333. Proceedings of Machine Learning Research. PMLR; 2026:794-833. https://proceedings.mlr.press/v333/lopez26a.html

23. Lindenmeyer A, Blattmann M, Franke S, Neumuth T, Schneider D. Towards Trustworthy AI in Healthcare: Epistemic Uncertainty Estimation for Clinical Decision Support. J Pers Med. 2025;15(2):58. doi:10.3390/jpm15020058

24. Tranchellini F, Farag Y, Jutzeler C, Meegahapola L. Evaluating deep learning sepsis prediction models in ICUs under distribution shift: a multi-centre retrospective cohort study. Npj Digit Med. 2026;9(1):306. doi:10.1038/s41746-026-02364-4

25. Ulmer D, Meijerink L, Cinà G. Trust Issues: Uncertainty Estimation Does Not Enable Reliable OOD Detection On Medical Tabular Data. In: Proceedings of the Machine Learning for Health NeurIPS Workshop. Vol 136. Proceedings of Machine Learning Research. PMLR; 2020:341-354.

26. Ren J, Fort S, Liu J, Roy AG, Padhy S, Lakshminarayanan B. A Simple Fix to Mahalanobis Distance for Improving Near-OOD Detection. *arXiv*. Preprint posted online June 16, 2021:arXiv:2106.09022. doi:10.48550/arXiv.2106.09022

27. Postels J, Segu M, Sun T, et al. On the Practicality of Deterministic Epistemic Uncertainty. arXiv. Preprint posted online July 5, 2022:arXiv:2107.00649. doi:10.48550/arXiv.2107.00649

28. Mukhoti J, Kirsch A, van Amersfoort J, Torr PHS, Gal Y. Deep Deterministic Uncertainty: A Simple Baseline. *arXiv*. Preprint posted online January 28, 2022:arXiv:2102.11582. doi:10.48550/arXiv.2102.11582

29. Frey N, Agha-Mir-Salim L, Hinz E, Poncette AS, Balzer F. Assessing Healthcare Stakeholder Understanding of Machine Learning Documentation. In: Andrikopoulou E, Gallos P, Arvanitis TN, et al., eds. Studies in Health Technology and Informatics. IOS Press; 2025. doi:10.3233/SHTI250336

30. Agha-Mir-Salim L, Mosch L, Klopfenstein SAI, et al. Artificial Intelligence Competencies in Postgraduate Medical Training in Germany. In: Séroussi B, Weber P, Dhombres F, et al., eds. Studies in Health Technology and Informatics. IOS Press; 2022. doi:10.3233/SHTI220589

31. Finlayson SG, Subbaswamy A, Singh K, et al. The Clinician and Dataset Shift in Artificial Intelligence. N Engl J Med. 2021;385(3):283-286. doi:10.1056/NEJMc2104626

32. Kohler S, Meyer-Eschenbach F, Michelena X, Frey N, Marschollek M, Eils R. Global Adoption of openEHR Clinical Data Repositories: A Vendor and Community Survey. medRxiv. Preprint posted online August 31, 2026. doi:10.64898/2026.08.27.26361529

33. Nestor B, McDermott MBA, Boag W, et al. Feature Robustness in Non-stationary Health Records: Caveats to Deployable Model Performance in Common Clinical Machine Learning Tasks. In: Proceedings of the 4th Machine Learning for Healthcare Conference. Vol 106. Proceedings of Machine Learning Research. PMLR; 2019:381-405. https://proceedings.mlr.press/v106/nestor19a.html

34. Agha-Mir-Salim L, Fehlhaber AL, Frey N, Krumpal I, Näher AF. Addressing Algorithmic Bias Through Data Disclosure: Evidence from a Randomized Experiment Among Physicians. SSRN. Preprint posted online 2026. doi:10.2139/ssrn.6328860

35. Liu JZ, Lin Z, Padhy S, Tran D, Bedrax-Weiss T, Lakshminarayanan B. Simple and Principled Uncertainty Estimation with Deterministic Deep Learning via Distance Awareness. *arXiv*. Preprint posted online October 26, 2020:arXiv:2006.10108. doi:10.48550/arXiv.2006.10108

36. van Amersfoort J, Smith L, Teh YW, Gal Y. Uncertainty Estimation Using a Single Deep Deterministic Neural Network. *arXiv*. Preprint posted online June 29, 2020:arXiv:2003.02037. doi:10.48550/arXiv.2003.02037

37. Johnson AEW, Bulgarelli L, Shen L, et al. MIMIC-IV, a freely accessible electronic health record dataset. Sci Data. 2023;10(1):1. doi:10.1038/s41597-022-01899-x

38. Pollard TJ, Johnson AEW, Raffa JD, Celi LA, Mark RG, Badawi O. The eICU Collaborative Research Database, a freely available multi-center database for critical care research. Sci Data. 2018;5(1):180178. doi:10.1038/sdata.2018.178

39. Harutyunyan H, Khachatrian H, Kale DC, Ver Steeg G, Galstyan A. Multitask learning and benchmarking with clinical time series data. Sci Data. 2019;6(1):96. doi:10.1038/s41597-019-0103-9

40. Wang S, McDermott MBA, Chauhan G, Ghassemi M, Hughes MC, Naumann T. MIMIC-Extract: a data extraction, preprocessing, and representation pipeline for MIMIC-III. In: *Proceedings of the ACM Conference on Health*, Inference, and Learning. ACM; 2020:222–235. doi:10.1145/3368555.3384469

41. Giesa N, Zhumagambetov R, Sekutowitcz M, et al. Benchmarking imputation methods on real-world clinical time series with simulated spatio-temporal missingness. Nat Commun. 2026;17(1):10491. doi:10.1038/s41467-026-77303-8

42. Carrasco-Ribelles LA, Llanes-Jurado J, Gallego-Moll C, et al. Prediction models using artificial intelligence and longitudinal data from electronic health records: a systematic methodological review. J Am Med Inform Assoc. 2023;30(12):2072–2082. doi:10.1093/jamia/ocad168

43. Hama T, Alsaleh MM, Allery F, et al. Enhancing Patient Outcome Prediction Through Deep Learning With Sequential Diagnosis Codes From Structured Electronic Health Record Data: Systematic Review. J Med Internet Res. 2025;27:e57358. doi:10.2196/57358

44. Lipton ZC, Kale DC, Wetzel R. Modeling Missing Data in Clinical Time Series with RNNs. *arXiv*. Preprint posted online November 11, 2016:arXiv:1606.04130. doi:10.48550/arXiv.1606.04130

45. Che Z, Purushotham S, Cho K, Sontag D, Liu Y. Recurrent Neural Networks for Multivariate Time Series with Missing Values. Sci Rep. 2018;8(1):6085. doi:10.1038/s41598-018-24271-9

46. Vaswani A, Shazeer N, Parmar N, et al. Attention Is All You Need. *arXiv*. Preprint posted online June 12, 2017:arXiv:1706.03762. doi:10.48550/arXiv.1706.03762

47. Miyato T, Kataoka T, Koyama M, Yoshida Y. Spectral Normalization for Generative Adversarial Networks. *arXiv*. Preprint posted online February 16, 2018:arXiv:1802.05957. doi:10.48550/arXiv.1802.05957

48. Kingma DP, Ba J. Adam: A Method for Stochastic Optimization. *arXiv*. Preprint posted online January 30, 2017:arXiv:1412.6980. doi:10.48550/arXiv.1412.6980

49. Akiba T, Sano S, Yanase T, Ohta T, Koyama M. Optuna: A Next-generation Hyperparameter Optimization Framework. *arXiv*. Preprint posted online July 25, 2019:arXiv:1907.10902. doi:10.48550/arXiv.1907.10902

50. Lakshminarayanan B, Pritzel A, Blundell C. Simple and Scalable Predictive Uncertainty Estimation using Deep Ensembles. *arXiv*. Preprint posted online November 4, 2017:arXiv:1612.01474. doi:10.48550/arXiv.1612.01474

51. Gal Y, Ghahramani Z. Dropout as a Bayesian Approximation: Representing Model Uncertainty in Deep Learning. In: Proceedings of The 33rd International Conference on Machine Learning. Vol 48. Proceedings of Machine Learning Research. PMLR; 2016:1050-1059. https://proceedings.mlr.press/v48/gal16.html

52. Smith L, Gal Y. Understanding Measures of Uncertainty for Adversarial Example Detection. *arXiv*. Preprint posted online March 22, 2018:arXiv:1803.08533. doi:10.48550/arXiv.1803.08533

53. Sun Y, Ming Y, Zhu X, Li Y. Out-of-Distribution Detection with Deep Nearest Neighbors. arXiv. Preprint posted online December 8, 2022:arXiv:2204.06507. doi:10.48550/arXiv.2204.06507

54. Van Calster B, Nieboer D, Vergouwe Y, De Cock B, Pencina MJ, Steyerberg EW. A calibration hierarchy for risk models was defined: from utopia to empirical data. J Clin Epidemiol. 2016;74:167–176. doi:10.1016/j.jclinepi.2015.12.005

55. Pakdaman Naeini M, Cooper G, Hauskrecht M. Obtaining Well Calibrated Probabilities Using Bayesian Binning. Proc AAAI Conf Artif Intell. 2015;29(1). doi:10.1609/aaai.v29i1.9602

56. Nixon J, Dusenberry M, Jerfel G, et al. Measuring Calibration in Deep Learning. *arXiv*. Preprint posted online August 7, 2020:arXiv:1904.01685. doi:10.48550/arXiv.1904.01685

57. El-Yaniv R, Wiener Y. On the Foundations of Noise-free Selective Classification. J Mach Learn Res. 2010;11:1605–1641.

58. Geifman Y, El-Yaniv R. Selective Classification for Deep Neural Networks. *arXiv*. Preprint posted online June 1, 2017:arXiv:1705.08500. doi:10.48550/arXiv.1705.08500

59. Geifman Y, Uziel G, El-Yaniv R. Bias-Reduced Uncertainty Estimation for Deep Neural Classifiers. *arXiv*. Preprint posted online April 24, 2019:arXiv:1805.08206. doi:10.48550/arXiv.1805.08206

