## Supplementary Information for "Evaluating uncertainty for detecting performance loss and guiding deferral after model transfer between hospital systems"

#### Supplementary Note 1 Formal definitions

This section gives the equations for the quantities that the Methods describe in words.

##### Notation

Patients (ICU stays) are indexed by $i$. For patient $i$, a model returns the log-odds $z_i$ and the predicted probability of in-hospital death $p_i = \sigma(z_i)$, where $\sigma(z) = 1/(1 + \exp(-z))$ is the logistic function; $y_i \in \{0, 1\}$ is the observed outcome. $n$ is the number of patients in the cohort that is evaluated, and $\pi$ is the mortality rate of this cohort, the mean of the $y_i$. Each architecture was trained with $S = 10$ seeds, indexed by $s$, and Monte Carlo dropout used $T = 30$ passes, indexed by $t$.

##### Inputs and classification threshold

The twelve continuous variables were standardized. With $x_{ij}$ a value of variable $j$ for patient $i$, and $\mu_j^{\text{train}}$ and $\sigma_j^{\text{train}}$ the mean and standard deviation of variable $j$ in the MIMIC-IV training data, the same transformation was applied to both databases:

$$\tilde{x}_{ij} = \frac{x_{ij} - \mu_j^{\text{train}}}{\sigma_j^{\text{train}}} \qquad (S1)$$

A prediction was counted as positive if it exceeded the threshold $\tau$, the mortality rate of the training pool. The error label $e_i$ marks a prediction that differs from the observed outcome:

$$\tau = 0.1288, \quad \hat{y}_i = 1\{p_i > \tau\}, \quad e_i = 1\{\hat{y}_i \neq y_i\} \qquad (S2)$$

##### Encoder constraints

In the constrained variants, every input and recurrent weight matrix $W$ of the GRU or the LSTM was replaced in the forward pass by

$$W_{\text{SN}} = \frac{W}{\max(1, \hat{\sigma}(W)/c)} \qquad (S3)$$

where $\hat{\sigma}(W)$ is an estimate of the largest singular value of $W$, updated by one step of power iteration per forward pass during training and held fixed at evaluation[1]. With $c = 1$, a matrix is rescaled only if this estimate exceeds 1. The prediction head was not normalized.

The residual block maps the final state $h$ of the recurrent encoder to the representation that enters the prediction head,

$$h_{\text{feat}} = h + C_R g(h), \quad g(h) = W_2 \text{ReLU}(W_1 h) \qquad (S4)$$

with $W_1$ and $W_2$ normalized as in Eq. S3 and $C_R = 0.9$. The resulting bounds on distances are given in Supplementary Note 3.

#### Training loss

All models were trained with the binary cross-entropy, which for one patient is

$$\ell_i = -[y_i \ln(p_i) + (1 - y_i)\ln(1 - p_i)] \quad (S5)$$

#### Deep ensembles and Monte Carlo dropout

The deep ensemble of an architecture averages the predicted probabilities $p_i^{(s)}$ of its ten seed models,

$$\overline{p}_i = \frac{1}{S}\sum_{s=1}^{S} p_i^{(s)} \quad (S6)$$

For deep ensembles, these ten probabilities are the repeated predictions $p_i^{(t)}$ in Eq. 2 to 4, with T = 10. For Monte Carlo dropout, pass t, with every dropout layer that was active in training switched on, returns the log-odds $z_i^{(t)}$, and the repeated predictions are $p_i^{(t)} = \sigma\left(z_i^{(t)}\right)$, with T = 30.

The logit variance is the variance of the log-odds across the passes, computed before the logistic function is applied:

$$V_{\mathrm{epi},i} = \frac{1}{T}\sum_{t=1}^{T}\left(z_i^{(t)} - \overline{z}_i\right)^2, \quad \overline{z}_i = \frac{1}{T}\sum_{t=1}^{T} z_i^{(t)} \quad (S7)$$

#### Discrimination and calibration

For a score $u$ and two groups of patients, a positive group with $n_1$ patients and a negative group with $n_0$ patients, the area under the receiver operating characteristic curve is

$$\mathrm{AUROC} = \frac{1}{n_1 n_0}\sum_{j\in\mathrm{pos}}\sum_{i\in\mathrm{neg}}\left[1\{u_j > u_i\} + \frac{1}{2}1\{u_j = u_i\}\right] \quad (S8)$$

It is the probability that a randomly drawn patient of the positive group has a higher score than a randomly drawn patient of the negative group, with ties counted as one half. For discrimination, the score is $p_i$, the positive group consists of the patients who died, and the negative group of the survivors.

The area under the precision–recall curve (AUPRC) was computed as average precision and is reported relative to the mortality rate of the cohort,

$$\mathrm{Lift} = \frac{\mathrm{AUPRC}}{\pi}. \quad (S9)$$

The calibration slope $b$ and the intercept $a$ are the coefficients of the logistic recalibration model

$$\text{logit P}(Y_i = 1 \mid z_i) = a + bz_i \qquad (S10)$$

For the expected calibration error, patients were grouped into ten bins $q$ of predicted probability (Supplementary Note 6). With $n_q$ patients, observed mortality $r_q$ and mean predicted probability $p_q$ in bin $q$, the error is the sum over the non-empty bins,

$$\text{ECE} = \sum_q \frac{n_q}{n} |r_q - p_q| \qquad (S11)$$

The cross-entropy of a cohort is the mean of Eq. S5 over its patients,

$$\text{CE} = \frac{1}{n} \sum_{i=1}^{n} \ell_i \qquad (S12)$$

and skill compares it with $\text{CE}_0$, the cross-entropy of a constant prediction of $\pi$ (Eq. S19):

$$\text{Skill} = 1 - \frac{\text{CE}}{\text{CE}_0} \qquad (S13)$$

### Supplementary Note 2 Sensitivity analysis: classification threshold

All error labels in the main text use the operating threshold $\tau = 0.1288$, the event rate of the training pool, with a prediction counted as positive if $p > \tau$. As a sensitivity analysis results were recomputed at $\tau = 0.5$; they are reported alongside in Supplementary Table 8.

At $\tau = 0.5$, every uncertainty measure identified errors less well. Within each configuration, predictive entropies still ranked above mutual information, mutual information above the class-conditional density, and the logit-variance component remained inverted. The Monte Carlo entropy and the single-model entropy stayed within 0.01 of each other at both thresholds (0.761 vs 0.755, 0.753 vs 0.744, and 0.770 vs 0.767 at $\tau = 0.5$), and the density on L2-normalized representations exceeded the single-model entropy for the GRU (0.764 vs 0.755).

### Supplementary Note 3 Encoder constraints

Spectral normalization (Eq. S3) rescales every recurrent weight matrix by its largest singular value, estimated by one step of power iteration per forward pass, with bound c = 1; this bounds the growth of distances through the encoder from above. The residual block (Eq. S4) adds a spectrally normalized branch scaled by $C_R$= 0.9 to the identity, so that, across this block alone, distances in $h_{feat}$ are bounded below by a factor of $1 - C_R = 0.1$ and above by $1 + C_R = 1.9$ of the distances in $h$, its input. This is the bi-Lipschitz property that distance-aware methods require [1,2], but it holds only for the final block: for the recurrent encoder that precedes it, spectral normalization gives an upper bound and no lower bound, so no distance preservation from the patient input to the representation is guaranteed end to end.

#### Supplementary Note 4 Training details

Hyperparameters were selected per configuration with Optuna[3] using a seeded tree-structured Parzen estimator[4] and a median pruner, 30 trials per architecture, minimizing the validation loss of a single deterministic pass; the constrained variants used the values of their base architecture (search space: Supplementary Table 3; selected values: Supplementary Table 4). Models were trained with Adam for a fixed number of epochs without early stopping. GPU computations were run in deterministic mode, so that a training seed fully determines the resulting model. Monte Carlo dropout masks were drawn with a separate fixed seed, so that ensemble members differ only in training and not in mask sampling.

#### Supplementary Note 5 Calibration measures and temperature scaling

Calibration-in-the-large is the intercept a of the recalibration model with the slope fixed at one,

$$\text{logit}\,\mathrm{P}(Y_i = 1 \mid z_i) = a_0 + z_i \qquad (S14)$$

with $z_i$ the logit of the predicted probability. The per-decile ratio of observed to predicted risk,

$$f_q = \frac{\overline{r}_q}{\overline{p}_q}, \qquad \overline{p}_q > 0 \qquad (S15)$$

shows where miscalibration occurs ($f_q > 1$: risk underestimated in bin $q$).

We examined whether the lower calibration slopes after transfer were consistent with the reduced separation between survivors and non-survivors. Predictions were compared on the log-odds scale. Their pooled variability within the two outcome groups remained almost unchanged (ratio of within-group standard deviations, eICU-CRD to MIMIC-IV: 0.98–1.00). However, the difference in mean log-odds between non-survivors and survivors decreased from 2.4–2.7 in MIMIC-IV to 1.7–1.9 in eICU-CRD.

Under the binormal model, if within-group variability remains unchanged, a reduction in group separation alone would imply a proportional decrease in calibration slope. Using the AUROC to quantify this separation, the expected transfer slope is:

$$b_{\text{transfer}} \approx b_{\text{in-domain}} \frac{\Phi^{-1}(\text{AUROC}_{\text{transfer}})}{\Phi^{-1}(\text{AUROC}_{\text{in-domain}})} \qquad (S16)$$

where (b) denotes the calibration slope and $\Phi^{-1}$ the inverse standard normal cumulative distribution function. Expected and observed transfer slopes differed by no more than 0.03 across all three base architectures.

As a sensitivity analysis, a straight line was fitted through the ten decile points of Fig. 1 (logit of the observed mortality against the mean log-odds of each decile, weighted by the number of patients per decile, deciles without deaths excluded). This binned slope was 0.90–1.09 on the internal test set and 0.47–0.61 on the external cohort, against 0.84–0.96 and 0.57–0.64 for the logistic recalibration; both methods show the fall after transfer.

Temperature scaling divides every logit by a single temperature $\theta$ fitted by minimizing the negative log-likelihood on the in-domain validation set,

$$p_{i,\text{cal}} = \sigma\left(\frac{z_i}{\theta}\right), \qquad \theta > 0 \tag{S17}$$

and changes the calibration slope by exactly the factor $\theta$,

$$b\left(\frac{z}{\theta}\right) = \theta\, b(z) \tag{S18}$$

so that a temperature fitted in-domain changes the transfer slope in a way that can be read off directly. Because temperature scaling is monotone, it leaves every rank-based measure unchanged (AUROC, AUPRC, and all Level 1–3 results of entropy). The skill score of the main text compares the cross-entropy with that of a constant prediction of the cohort prevalence $\pi$,

$$CE_0 = -[\pi \ln(\pi) + (1 - \pi) \ln(1 - \pi)] \tag{S19}$$

Results for all seven configurations, before and after temperature scaling: Supplementary Table 5.

### Supplementary Note 6 Binning of predicted probabilities

All binned quantities, the expected calibration error, the per-decile ratios (Eq. S15) and the decile points in Fig. 1, use the same rule[5,6]. Within each model configuration and cohort, the raw predictions of the ten seed models were pooled and their deciles taken as bin edges; the same numerical edges were applied to every seed and to both the raw and the temperature-scaled predictions, so that patients can change bins after scaling. Edges were computed separately for the two cohorts. Binned calibration measures depend on the choice of bins [7]; equal-count bins were chosen because the outcome is rare (69% to 78% of predictions below 0.1), and the highest decile therefore pools all predictions above roughly 0.3. The calibration analysis was repeated with ten equal-width bins over the full scale, with bin occupancies and 95% Wilson intervals computed per seed model and averaged (Supplementary Fig. 5), and with a flexible spline curve (Fig. 1).

### Supplementary Note 7 Density variants and fitting set

Besides the class-conditional Gaussian of the main text (Eq. 5 of the main text), four definitions of representation density were evaluated on the same representations and fitting set: a covariance shared across classes, scored as the mixture log-density; a per-class Gaussian mixture with three components; the class-conditional Gaussian on $L_2$-normalized representations, following the feature normalization of nearest-neighbour detection[8], with the norm floored at $10^{-12}$; and a background-corrected relative density score, the negative log-density of the class mixture minus that of a single background Gaussian over all training representations. The original relative Mahalanobis distance of Ren et al.[9], with a covariance $\Sigma$ pooled within classes and a background Gaussian $(\mu_0, \Sigma_0)$, was computed as a declared addition,

$$s_{\text{RMD}}(h) = \min_c[(h - \mu_c)^{\mathrm{T}}\Sigma^{-1}(h - \mu_c) - (h - \mu_0)^{\mathrm{T}}\Sigma_0^{-1}(h - \mu_0)] \tag{S20}$$

As a sensitivity analysis, the density was refitted on a class-balanced subset of the training set, all 3,780 stays with the outcome and 3,780 stays without it drawn without replacement with a fixed seed, with class priors of one half each; cohort separation changed by less than 0.01 in every configuration.

#### Supplementary Note 8 Selective prediction: details

The oracle area in the excess area (Eq. 6 of the main text) is computed with the same finite sum as the area itself, applied to the error labels sorted with all correct predictions first. Patients were ranked by a stable sort, so that patients with identical scores are accepted in the order of the cohort file; identical scores arise for stays with identical imputed inputs. Risk at 80% coverage is $R(\lfloor 0.8n \rfloor)$. Levels 2 and 3 were computed for both cohorts; the main text reports the external cohort and uses the internal test set as a baseline (Supplementary Table 12). The misclassification rate at 80% coverage is not directly comparable between cohorts, because the fixed classification threshold yields a different mix of false alarms and missed deaths; the excess area, which averages over all deferral budgets, is the comparable quantity.

#### Supplementary Note 9 Composition analysis and associations

To separate a shift in mutual information from a shift in the distribution of predictions, patients of both cohorts were assigned to ten deciles $q$ of the deterministic model's predicted probability, with edges from both cohorts and all ten seeds pooled. With $\bar{m}_{c,q}$ the mean mutual information and $w_{c,q}$ the share of patients in decile $q$ of cohort $c$ (internal, in; external, ex), the difference between the cohorts at common weights is

$$\Delta_{within} = \sum_q \frac{1}{2}\left(w_{in,q} + w_{ex,q}\right)\left(\bar{m}_{ex,q} - \bar{m}_{in,q}\right) \qquad (S21)$$

where deciles empty in one cohort contribute no term. This is a descriptive standardization, not a causal attribution. At common weights, the mean difference between the cohorts changed from −0.00062 to +0.00008 nats for the GRU and from −0.00021 to −0.00002 nats for the transformer; for the LSTM it changed sign, from −0.00036 to +0.00037 nats.

Associations between density measures and predictive entropy (Supplementary Table 7) are patient-level Spearman correlations on the external cohort, per configuration and seed, summarized as the mean with the range over seeds; $|\rho| \geq 0.70$ was read as a measure that largely tracks the prediction and $|\rho| < 0.40$ as largely independent of it. In a post hoc check of why the logit-variance component was inverted, it correlated negatively with the Monte Carlo predictive entropy of the same model (Spearman $\rho$ −0.32, −0.46, and −0.79 for the GRU, LSTM, and transformer; seed intervals −0.42 to −0.23, −0.52 to −0.40, and −0.84 to −0.75) and positively with the absolute log-odds (0.35, 0.51, and 0.81). Among the tenth of external patients with the largest logit variance, 13.0%, 14.8%, and 3.4% were misclassified, against 49.1%, 54.0%, and 62.4% among the tenth with the smallest.

#### Supplementary Note 10 Training length without early stopping

Two post hoc analyses addressed the fixed training length without early stopping (Supplementary Note 4). First, of the 70 trained models (seven model sets, ten seeds each), 66 had reached their lowest validation loss before the final epoch, and at the end of training the loss was on average 0.5% to 2.3% above this minimum, depending on architecture and variant (Supplementary Table 10, Supplementary Fig. 9); this is within the range at which established early-stopping rules end training (1% to 5% above the minimum[10,11]). Second, each model of the three base architectures was then assessed at the epoch of its lowest validation loss, by repeating the deterministic training up to that epoch. The AUROC then changed by at most 0.006 and its loss after transfer remained close to 0.10. The calibration slope improved, from 0.84–0.96 to 0.92–1.02 on the internal test set and from 0.57–0.64 to 0.64–0.68 on the external cohort, and still fell by about one third after transfer. The predictive entropy separated the cohorts at 0.464, 0.452, and 0.454 instead of 0.475, 0.466, and 0.462, below 0.5 in every seed; error detection changed by at most 0.007 and the E-AURC by at most 0.002 (Supplementary Table 11). All conclusions were unchanged, and we therefore consider the models to be without substantial overtraining for the purposes of this study.

### Supplementary Figures

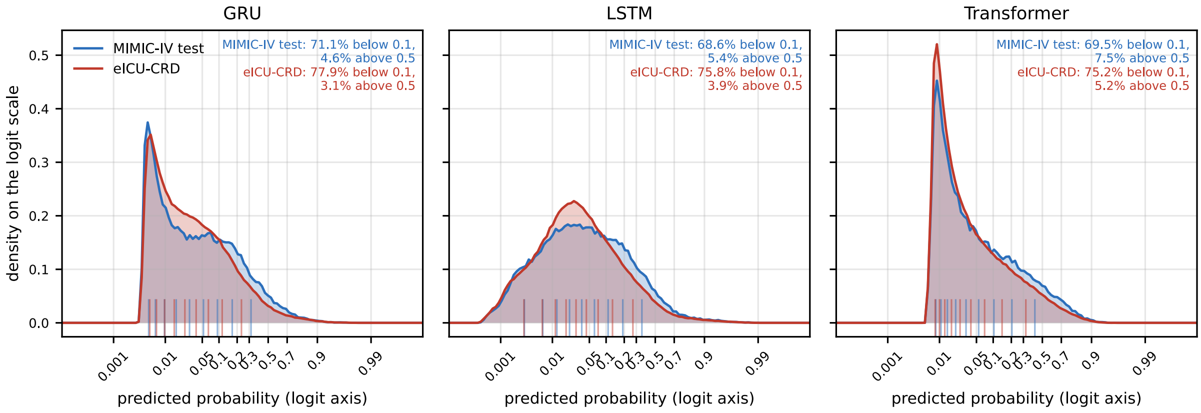


**Supplementary Fig. 1 | Distribution of predicted mortality probabilities in both cohorts.** Predictions of the ten seed models for each architecture on a logit axis, which resolves the many low predictions. Blue: internal test set (MIMIC-IV, 7,298 stays); red: external cohort (eICU-CRD, 66,934 stays). Percentages give the share of predictions below 0.1 and above 0.5. Short ticks at the bottom of each panel mark the decile edges of each cohort, as used for Fig. 1.

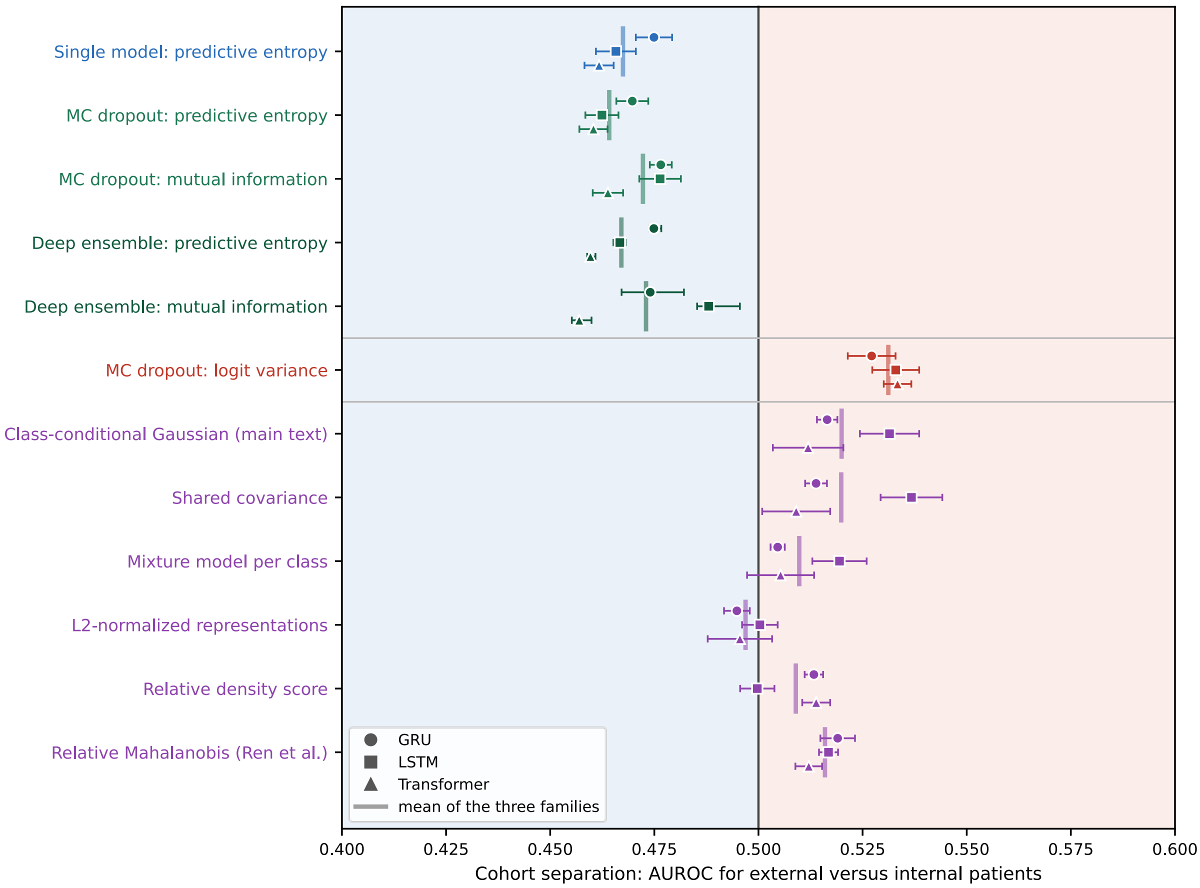


**Supplementary Fig. 2 | Discrimination between internal and external cohorts using uncertainty and representation-based scores.** AUROC >0.5 indicates higher scores in the external cohort; AUROC <0.5 indicates higher scores in the internal cohort. Symbols represent model architectures; vertical markers indicate the mean across architectures. Horizontal bars show the 95% t-interval across the ten seed models; for deep ensembles, the value of the full ensemble with the range over the ten nine-member ensembles. Blue background: AUROC below 0.5; red background: above 0.5. Label colors group the measures: blue, single model; green, Monte Carlo (MC) dropout and deep ensembles; red, logit variance; purple, representation density.

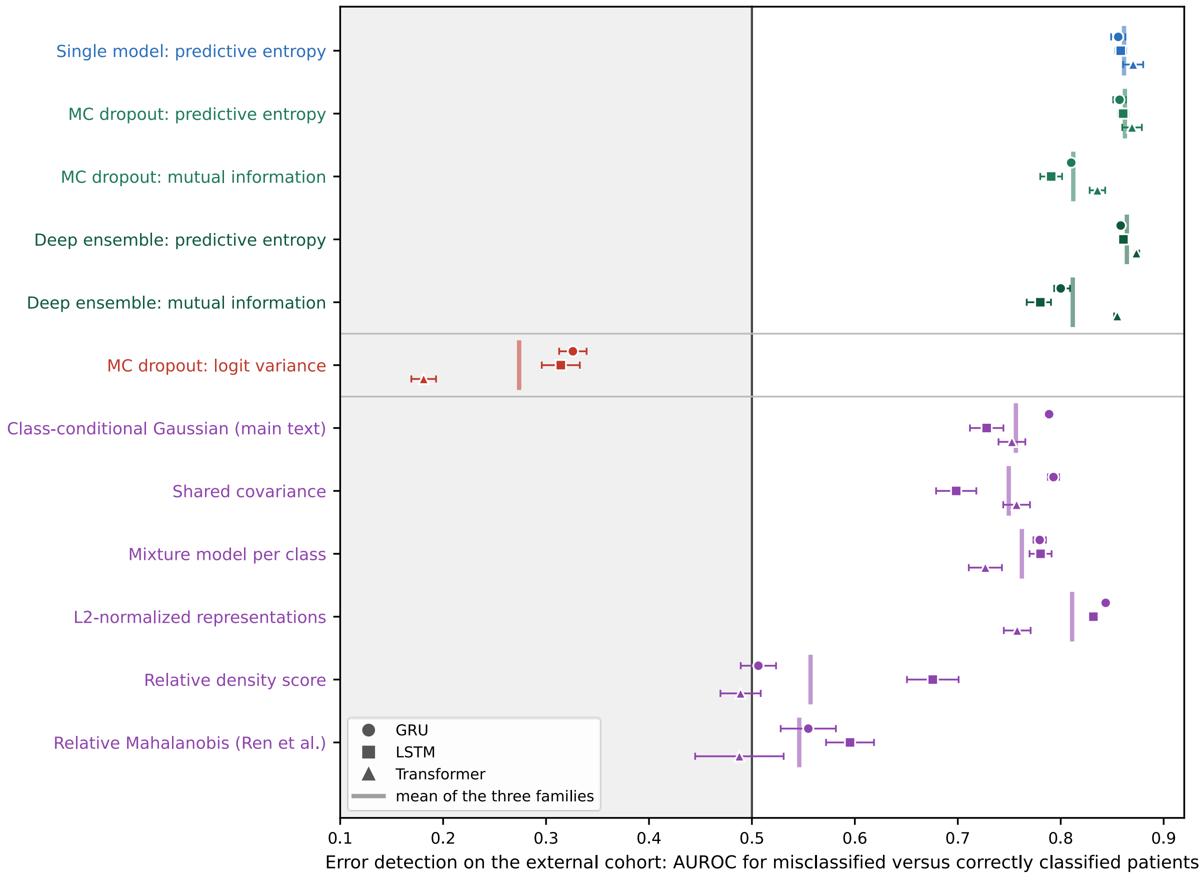


**Supplementary Fig. 3 | Error detection on the external cohort using uncertainty and representation-based scores**. AUROC >0.5 indicates higher scores for misclassified than correctly classified patients; 0.5 indicates chance-level discrimination. Symbols represent model architectures; vertical markers indicate the mean across architectures. Horizontal bars show the 95% t-interval across the ten seed models; for deep ensembles, the value of the full ensemble with the range over the ten nine-member ensembles. Gray background: below chance. Label colors as in Supplementary Fig. 2.

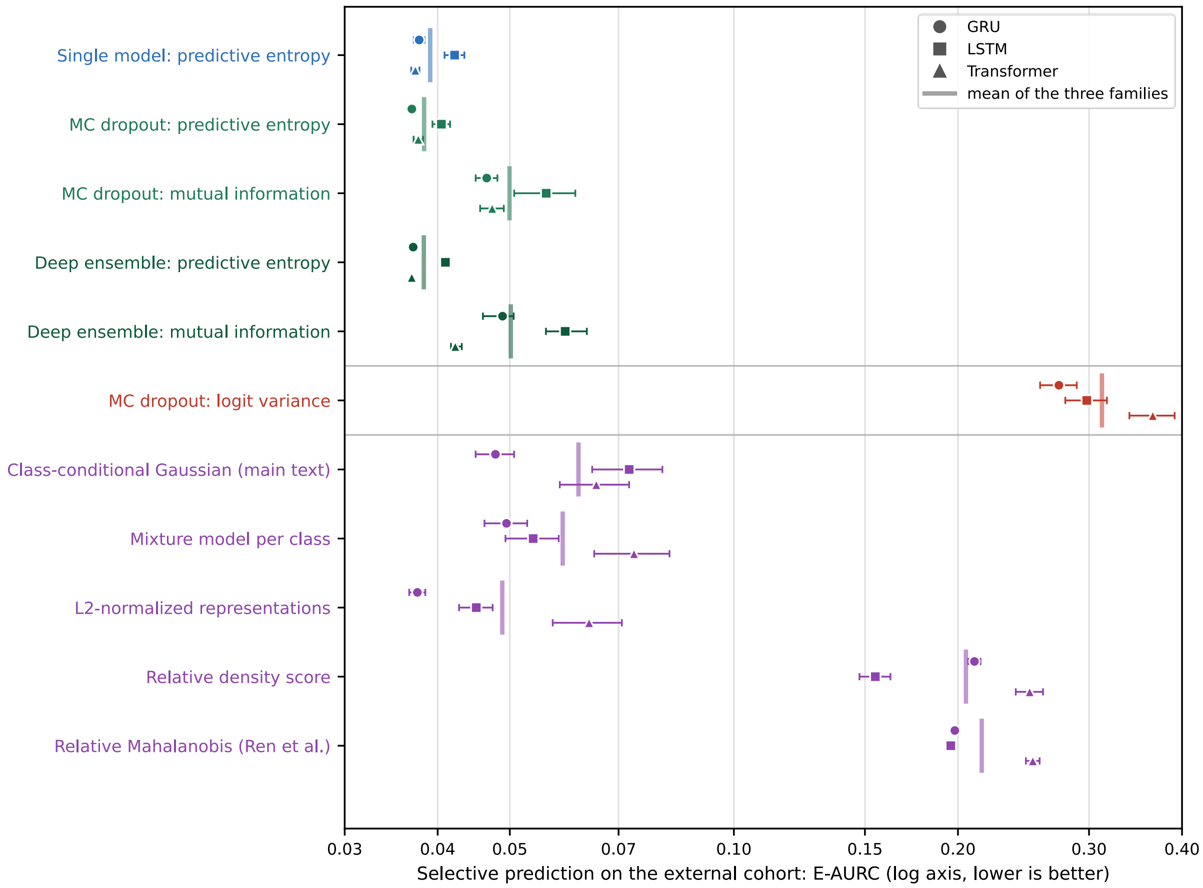


**Supplementary Fig. 4 | Selective prediction on the external cohort using uncertainty and representation-based scores.** Lower excess area under the risk–coverage curve (E-AURC) indicates better selective prediction. Symbols represent model architectures; vertical markers indicate the mean across architectures. The horizontal axis uses a logarithmic scale. Horizontal bars show the 95% t-interval across the ten seed models; for deep ensembles, the value of the full ensemble with the range over the ten nine-member ensembles. Label colors as in Supplementary Fig. 2.

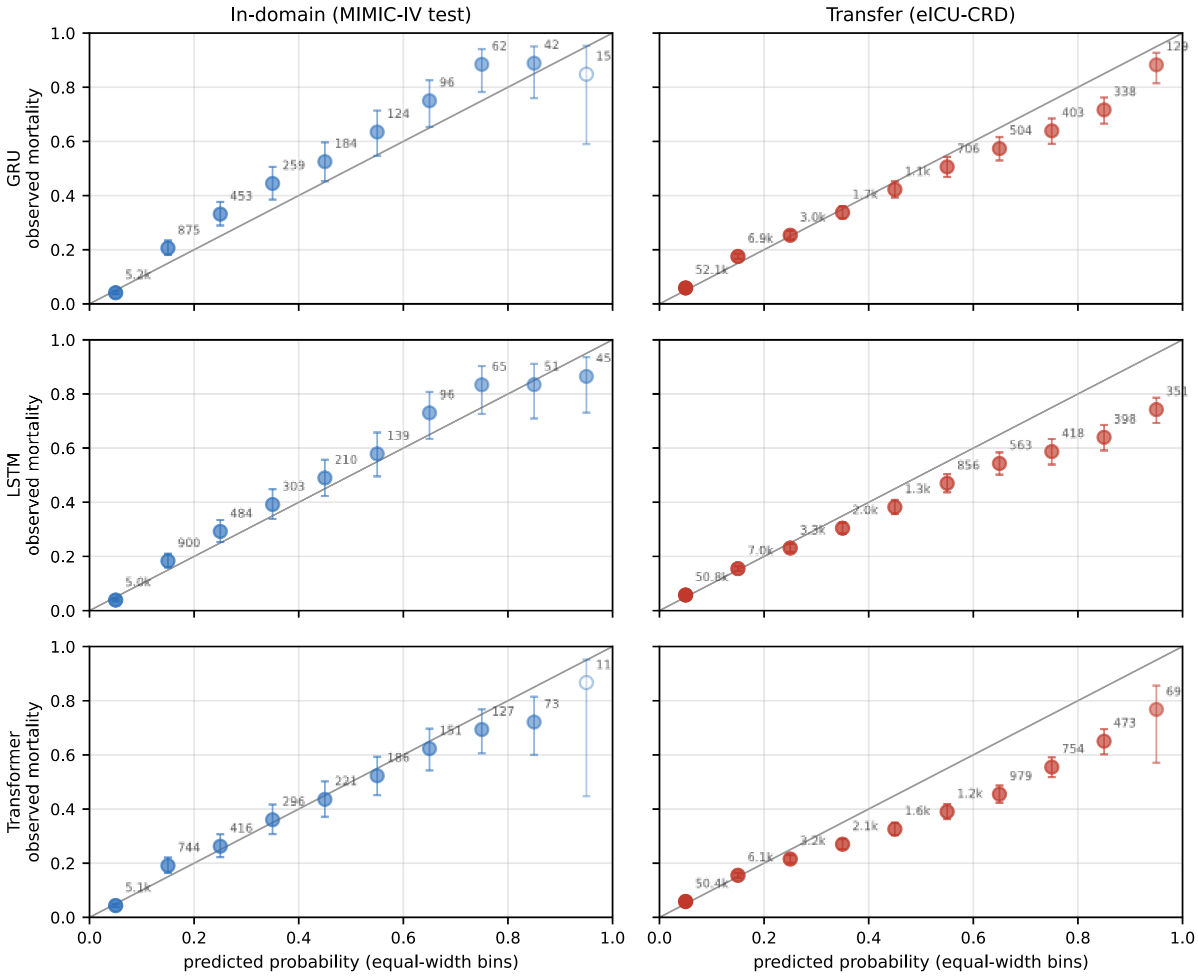


**Supplementary Fig. 5 | Calibration in ten equal-width bins of predicted probability.** Observed mortality at the bin midpoint for the internal test set (left, blue) and the external cohort (right, red), one row per architecture. Error bars show 95% Wilson intervals and numbers the patients per bin, both computed per seed model and averaged over the ten seeds. Open circles mark bins with fewer than 30 patients; the gray diagonal marks perfect calibration.

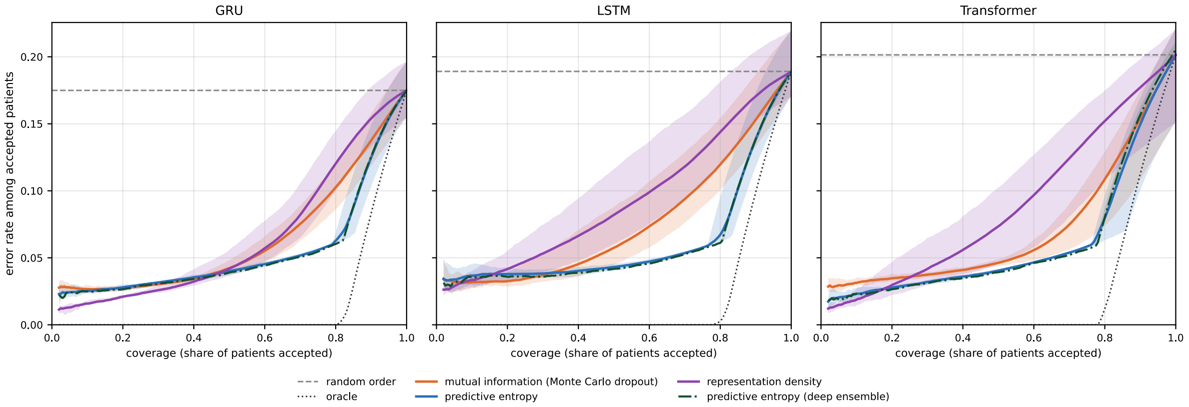


**Supplementary Fig. 6 | Risk–coverage curves on the external cohort**. Misclassification rate among the accepted patients against the share of patients accepted, with patients accepted from the most certain downward; one panel per architecture. Lines are means over the ten seed models, shaded bands the range over seeds; for the deep ensemble, the full ensemble with the range over the ten nine-member ensembles. The dashed line marks random order (the overall misclassification rate), the dotted line an oracle that accepts all correctly classified patients first.

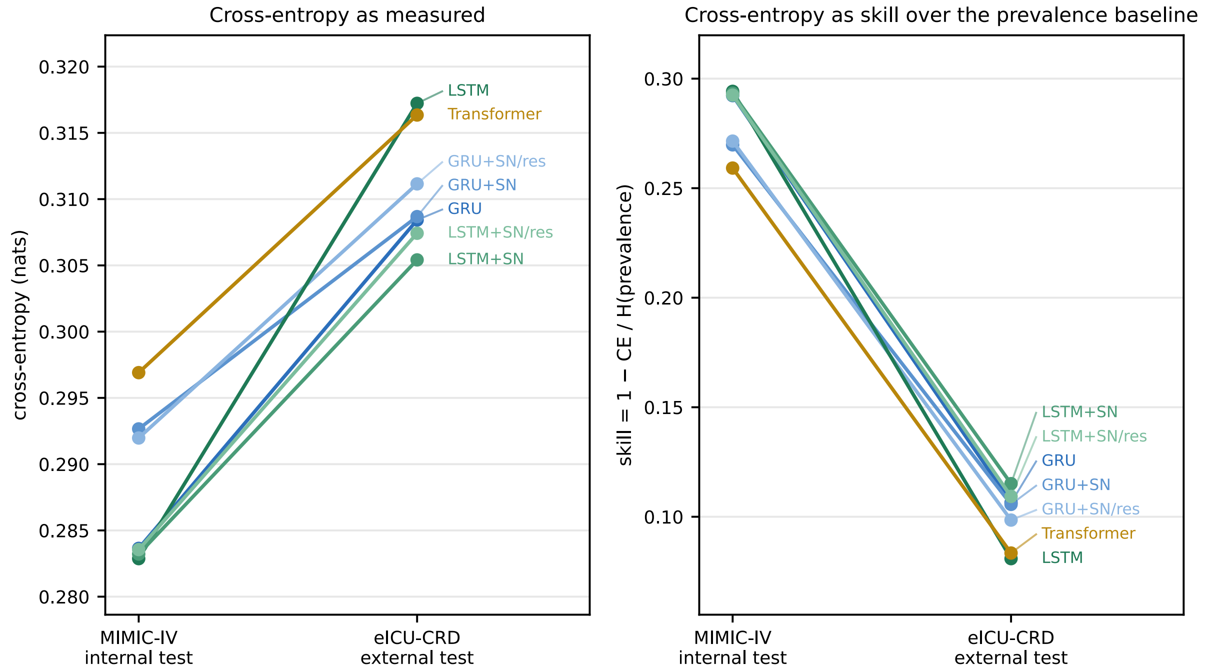


**Supplementary Fig. 7 | Cross-entropy of the predicted probabilities in the internal and the external test cohort (left) and skill relative to a constant prediction of the cohort prevalence (right).** After transfer, skill fell by 61% to 73% in every configuration.

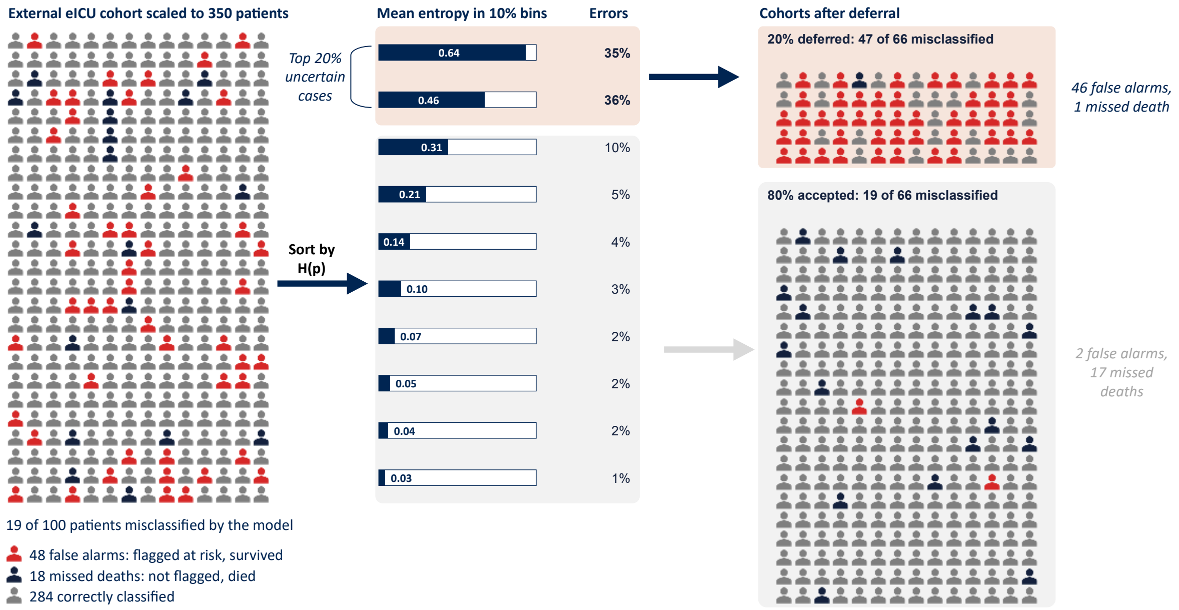


**Supplementary Fig. 8 | Deferral by predictive entropy with misclassifications separated by type.** External cohort scaled to 350 patients (single-model entropy, means over the three architectures and ten seeds). Errors: share of all misclassifications falling in the bin; a full bar corresponds to 0.69 nats.

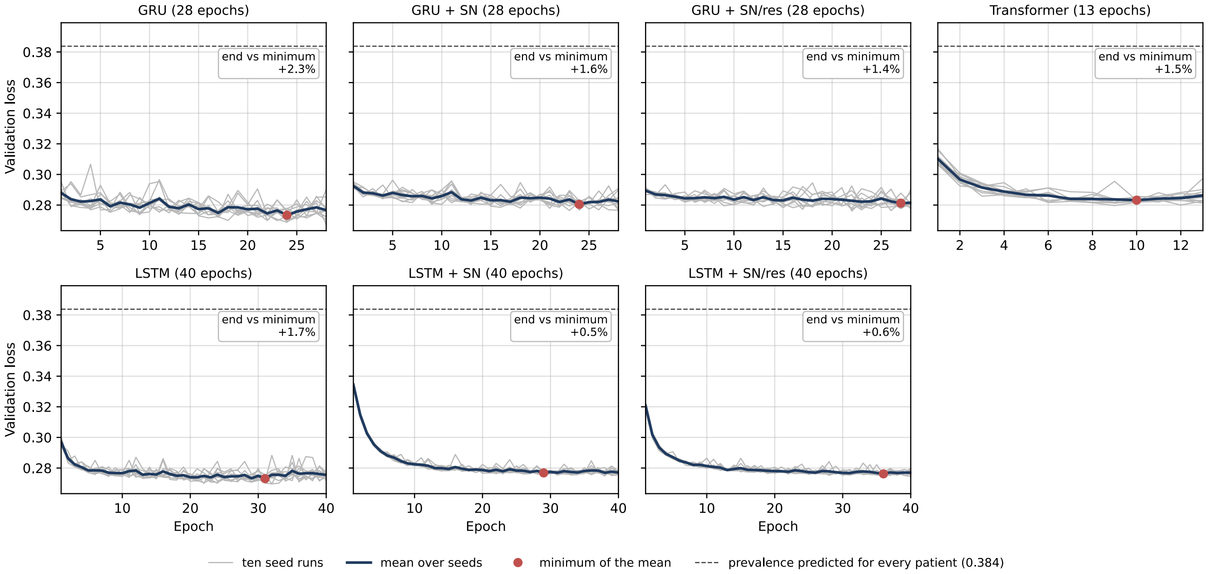


**Supplementary Fig. 9 | Validation loss per epoch for the seven model configurations.** One panel per configuration, with the number of training epochs in the panel title. Boxes give the mean increase of the last-epoch loss above each seed model's own minimum (Supplementary Table 10), so they can exceed the gap between the mean curve and its minimum. SN: spectral normalization; res: residual block.

### Supplementary Tables

**Supplementary Table 1 | Input variables, encoding and imputation**

| # | Variable | Unit | Clinical default (used only if never measured within 48 h) | Encoding | Channels |
|---|---|---|---|---|---|
| **1** | Heart rate | /min | 86 | standardized | 1 |
| **2** | Respiratory rate | /min | 19 | standardized | 1 |
| **3** | Systolic blood pressure | mmHg | 118 | standardized | 1 |
| **4** | Diastolic blood pressure | mmHg | 59 | standardized | 1 |
| **5** | Mean arterial pressure | mmHg | 77 | standardized | 1 |
| **6** | Oxygen saturation | % | 98 | standardized | 1 |
| **7** | Temperature | °C | 36.6 | standardized | 1 |
| **8** | Fraction of inspired oxygen | fraction | 0.21 | standardized | 1 |
| **9** | Glucose | mg/dL | 128 | standardized | 1 |
| **10** | pH | – | 7.4 | standardized | 1 |
| **11** | Weight | kg | 81 | standardized | 1 |
| **12** | Height | cm | 170 | standardized | 1 |
| **13** | Glasgow Coma Scale, total | points | 15 | one-hot, 13 levels (3–15) | 13 |
| | | | | **Total** | **25** |

Unlike the benchmark of Harutyunyan et al.[12], which takes the last measurement within an hour, imputes the normal value before the first measurement, and adds a binary mask per variable, we averaged measurements within an hour, filled hours before the first measurement with the first observed value, and used no masks. The same rules were applied to both databases.

**Supplementary Table 2 | Sensitivity analysis: external cohort restricted to admissions with a single ICU stay in the raw data**

| Quantity | Full external cohort (n = 66,934) | Strict subset (n = 56,556) |
|---|---|---|
| **Performance and calibration** | | |
| **AUROC, mortality** | 0.762 to 0.775 | 0.771 to 0.784 |
| **AUPRC lift** | 3.14 to 3.48 | 3.28 to 3.64 |
| **Calibration slope** | 0.569 to 0.698 | 0.589 to 0.720 |
| **ECE, raw** | 0.030 to 0.044 | 0.027 to 0.042 |
| **Skill** | 0.081 to 0.115 | 0.099 to 0.132 |
| **AUROC of the ensemble mean** | 0.767 to 0.777 | 0.776 to 0.786 |
| **Cohort separation (AUROC)** | | |
| **Single model: predictive entropy** | 0.428 to 0.475 | 0.426 to 0.474 |
| **MC dropout: predictive entropy** | 0.460 to 0.470 | 0.459 to 0.469 |
| **MC dropout: mutual information** | 0.464 to 0.477 | 0.463 to 0.476 |
| **Deep ensemble: predictive entropy** | 0.428 to 0.475 | 0.427 to 0.474 |
| **Deep ensemble: mutual information** | 0.448 to 0.488 | 0.446 to 0.487 |
| **Representation density (class-conditional Gaussian)** | 0.512 to 0.556 | 0.510 to 0.557 |
| **Error detection (AUROC)** | | |
| **Single model: predictive entropy** | 0.844 to 0.870 | 0.845 to 0.871 |
| **MC dropout: predictive entropy** | 0.857 to 0.869 | 0.859 to 0.870 |
| **MC dropout: mutual information** | 0.791 to 0.836 | 0.792 to 0.837 |
| **Deep ensemble: predictive entropy** | 0.845 to 0.874 | 0.846 to 0.874 |
| **Deep ensemble: mutual information** | 0.780 to 0.855 | 0.781 to 0.855 |
| **Representation density (class-conditional Gaussian)** | 0.728 to 0.789 | 0.729 to 0.791 |
| **Selective prediction (E-AURC)** | | |
| **Single model: predictive entropy** | 0.037 to 0.042 | 0.036 to 0.041 |
| **MC dropout: predictive entropy** | 0.037 to 0.040 | 0.036 to 0.040 |
| **MC dropout: mutual information** | 0.047 to 0.056 | 0.046 to 0.055 |
| **Deep ensemble: predictive entropy** | 0.037 to 0.041 | 0.036 to 0.040 |
| **Deep ensemble: mutual information** | 0.038 to 0.059 | 0.036 to 0.058 |
| **Representation density (class-conditional Gaussian)** | 0.048 to 0.072 | 0.047 to 0.071 |

Ranges across the seven configurations (three for Monte Carlo dropout) of the means over ten seed models; deep ensembles: values of the full ensemble. MC: Monte Carlo; E-AURC: excess area under the risk–coverage curve; ECE: expected calibration error.

**Supplementary Table 3 | Hyperparameter search space**

| Hyperparameter | Range / choices | Applies to |
|---|---|---|
| **learning rate** | 1e-5 … 1e-2 (log) | all |
| **num_layers** | 1 … 4 | all |
| **num_epochs** | 10 … 40 (fixed schedule, no early stopping) | all |
| **batch_size** | {32, 64, 128} | all |
| **dropout** | 0.2 … 0.8 | all |
| **hidden_size** | 32 … 256 | recurrent |
| **d_model** | {64, 128, 256} | transformer |
| **nhead** | {2, 4, 8} | transformer |
| **dim_feedforward** | 64 … 512 (step 64) | transformer |
| **optimizer / objective** | Adam; per-epoch validation BCE (one deterministic pass) | all |
| **search** | Optuna TPE (seeded); MedianPruner(10, 6) | all |

**Supplementary Table 4 | Selected hyperparameters per configuration**

| Configuration | learning_rate | num_layers | num_epochs | batch_size | dropout | hidden_size | d_model | nhead | dim_feedforward | n_trials |
|---|---|---|---|---|---|---|---|---|---|---|
| **GRU** | 2.83e-04 | 2 | 28 | 64 | 0.428 | 93 | - | - | - | 30 |
| **GRU + SN** | 2.83e-04 | 2 | 28 | 64 | 0.428 | 93 | - | - | - | |
| **GRU + SN/res** | 2.83e-04 | 2 | 28 | 64 | 0.428 | 93 | - | - | - | |
| **LSTM** | 5.46e-05 | 3 | 40 | 64 | 0.428 | 255 | - | - | - | 30 |
| **LSTM + SN** | 5.46e-05 | 3 | 40 | 64 | 0.428 | 255 | - | - | - | |
| **LSTM + SN/res** | 5.46e-05 | 3 | 40 | 64 | 0.428 | 255 | - | - | - | |
| **Transformer** | 6.60e-05 | 4 | 13 | 128 | 0.206 | - | 64 | 8 | 512 | 30 |

Constrained variants used the values of their base architecture (no separate search, therefore no trial count). SN: spectral normalization; res: residual block.

**Supplementary Table 5 | Calibration by configuration before and after temperature scaling**

| Configuration | Slope, MIMIC-IV | ECE raw, MIMIC-IV | ECE scaled, MIMIC-IV | Slope, eICU-CRD | ECE raw, eICU-CRD | ECE scaled, eICU-CRD | Temperature | Slope after scaling, eICU-CRD |
|---|---|---|---|---|---|---|---|---|
| GRU | 0.962 [0.935; 0.989] | 0.034 [0.027; 0.042] | 0.021 [0.018; 0.024] | 0.645 [0.631; 0.659] | 0.033 [0.030; 0.037] | 0.019 [0.017; 0.021] | 1.18 [1.12; 1.24] | 0.760 [0.730; 0.790] |
| GRU + SN | 1.057 [1.035; 1.078] | 0.041 [0.035; 0.048] | 0.030 [0.027; 0.033] | 0.698 [0.680; 0.716] | 0.036 [0.031; 0.041] | 0.021 [0.020; 0.022] | 1.16 [1.11; 1.21] | 0.807 [0.777; 0.837] |
| GRU + SN/res | 1.011 [0.987; 1.036] | 0.040 [0.035; 0.045] | 0.027 [0.024; 0.029] | 0.673 [0.658; 0.688] | 0.037 [0.032; 0.041] | 0.022 [0.021; 0.023] | 1.17 [1.12; 1.21] | 0.784 [0.760; 0.808] |
| LSTM | 0.915 [0.867; 0.964] | 0.022 [0.017; 0.028] | 0.012 [0.010; 0.015] | 0.569 [0.539; 0.599] | 0.034 [0.031; 0.038] | 0.027 [0.024; 0.030] | 1.13 [1.08; 1.18] | 0.641 [0.613; 0.668] |
| LSTM + SN | 1.044 [1.016; 1.073] | 0.023 [0.018; 0.029] | 0.019 [0.015; 0.022] | 0.656 [0.641; 0.671] | 0.030 [0.028; 0.032] | 0.026 [0.023; 0.028] | 1.05 [1.02; 1.09] | 0.690 [0.667; 0.713] |
| LSTM + SN/res | 1.038 [0.995; 1.081] | 0.026 [0.020; 0.031] | 0.020 [0.016; 0.023] | 0.648 [0.622; 0.674] | 0.031 [0.028; 0.033] | 0.025 [0.023; 0.027] | 1.07 [1.04; 1.11] | 0.693 [0.667; 0.720] |
| Transformer | 0.843 [0.812; 0.874] | 0.028 [0.020; 0.036] | 0.021 [0.015; 0.026] | 0.593 [0.573; 0.614] | 0.044 [0.040; 0.048] | 0.036 [0.030; 0.043] | 1.12 [1.06; 1.19] | 0.668 [0.613; 0.723] |

Means over ten seed models with the 95% t-interval across seeds in brackets. ECE: expected calibration error over deciles of predicted risk (Supplementary Note 6). SN: spectral normalization; res: residual block.

**Supplementary Table 6 | Complete results: all uncertainty measures, all levels, seven configurations**

| Uncertainty measure | Configuration | Cohort separation (AUROC) | Error detection (AUROC) | Selective prediction (E-AURC) |
|---|---|---|---|---|
| Single model: predictive entropy | GRU | 0.475 [0.471; 0.479] | 0.856 [0.849; 0.863] | 0.038 [0.037; 0.038] |
| Single model: predictive entropy | GRU + SN | 0.429 [0.427; 0.432] | 0.845 [0.837; 0.852] | 0.037 [0.037; 0.038] |
| Single model: predictive entropy | GRU + SN/res | 0.428 [0.426; 0.430] | 0.844 [0.837; 0.850] | 0.038 [0.037; 0.038] |
| Single model: predictive entropy | LSTM | 0.466 [0.461; 0.471] | 0.858 [0.853; 0.864] | 0.042 [0.041; 0.043] |
| Single model: predictive entropy | LSTM + SN | 0.437 [0.435; 0.439] | 0.863 [0.858; 0.868] | 0.040 [0.040; 0.041] |
| Single model: predictive entropy | LSTM + SN/res | 0.436 [0.434; 0.438] | 0.859 [0.854; 0.863] | 0.041 [0.040; 0.041] |
| Single model: predictive entropy | Transformer | 0.462 [0.458; 0.465] | 0.870 [0.861; 0.880] | 0.037 [0.037; 0.038] |
| MC dropout: predictive entropy (U_total) | GRU | 0.470 [0.466; 0.474] | 0.857 [0.851; 0.863] | 0.037 [0.036; 0.037] |
| MC dropout: predictive entropy (U_total) | LSTM | 0.462 [0.458; 0.466] | 0.861 [0.856; 0.866] | 0.040 [0.039; 0.042] |
| MC dropout: predictive entropy (U_total) | Transformer | 0.460 [0.457; 0.464] | 0.869 [0.860; 0.879] | 0.038 [0.037; 0.038] |
| MC dropout: aleatoric part (U_alea) | GRU | 0.470 [0.466; 0.473] | 0.858 [0.851; 0.864] | 0.037 [0.036; 0.037] |
| MC dropout: aleatoric part (U_alea) | LSTM | 0.462 [0.458; 0.466] | 0.861 [0.856; 0.866] | 0.040 [0.039; 0.042] |
| MC dropout: aleatoric part (U_alea) | Transformer | 0.460 [0.457; 0.464] | 0.869 [0.860; 0.879] | 0.038 [0.037; 0.038] |
| MC dropout: mutual information (MI) | GRU | 0.477 [0.474; 0.479] | 0.810 [0.806; 0.814] | 0.047 [0.045; 0.048] |
| MC dropout: mutual information (MI) | LSTM | 0.476 [0.471; 0.481] | 0.791 [0.780; 0.801] | 0.056 [0.051; 0.061] |
| MC dropout: mutual information (MI) | Transformer | 0.464 [0.460; 0.468] | 0.836 [0.828; 0.843] | 0.047 [0.046; 0.049] |
| MC dropout: logit variance (epistemic) | GRU | 0.527 [0.521; 0.533] | 0.326 [0.313; 0.339] | 0.273 [0.258; 0.289] |
| MC dropout: logit variance (epistemic) | LSTM | 0.533 [0.527; 0.539] | 0.314 [0.296; 0.333] | 0.298 [0.279; 0.317] |
| MC dropout: logit variance (epistemic) | Transformer | 0.533 [0.530; 0.537] | 0.181 [0.169; 0.193] | 0.365 [0.340; 0.391] |
| Deep ensemble: predictive entropy (U_total) | GRU | 0.475 [0.474; 0.477] | 0.858 [0.856; 0.860] | 0.037 [0.037; 0.037] |

| | | | | |
|---|---|---|---|---|
| Deep ensemble: predictive entropy (U_total) | GRU + SN | 0.429 [0.429; 0.430] | 0.846 [0.844; 0.848] | 0.037 [0.037; 0.037] |
| Deep ensemble: predictive entropy (U_total) | GRU + SN/res | 0.428 [0.428; 0.429] | 0.845 [0.843; 0.847] | 0.037 [0.037; 0.037] |
| Deep ensemble: predictive entropy (U_total) | LSTM | 0.467 [0.465; 0.468] | 0.861 [0.859; 0.862] | 0.041 [0.041; 0.041] |
| Deep ensemble: predictive entropy (U_total) | LSTM + SN | 0.437 [0.436; 0.437] | 0.864 [0.862; 0.865] | 0.040 [0.040; 0.040] |
| Deep ensemble: predictive entropy (U_total) | LSTM + SN/res | 0.437 [0.436; 0.437] | 0.859 [0.858; 0.860] | 0.040 [0.040; 0.041] |
| Deep ensemble: predictive entropy (U_total) | Transformer | 0.460 [0.459; 0.461] | 0.874 [0.872; 0.876] | 0.037 [0.037; 0.037] |
| Deep ensemble: aleatoric part (U_alea) | GRU | 0.475 [0.474; 0.477] | 0.858 [0.856; 0.860] | 0.037 [0.037; 0.037] |
| Deep ensemble: aleatoric part (U_alea) | GRU + SN | 0.429 [0.429; 0.430] | 0.846 [0.844; 0.848] | 0.037 [0.037; 0.037] |
| Deep ensemble: aleatoric part (U_alea) | GRU + SN/res | 0.428 [0.427; 0.429] | 0.845 [0.843; 0.847] | 0.037 [0.037; 0.037] |
| Deep ensemble: aleatoric part (U_alea) | LSTM | 0.466 [0.465; 0.467] | 0.861 [0.859; 0.862] | 0.041 [0.041; 0.041] |
| Deep ensemble: aleatoric part (U_alea) | LSTM + SN | 0.437 [0.436; 0.437] | 0.864 [0.862; 0.865] | 0.040 [0.040; 0.040] |
| Deep ensemble: aleatoric part (U_alea) | LSTM + SN/res | 0.436 [0.436; 0.437] | 0.859 [0.858; 0.860] | 0.040 [0.040; 0.041] |
| Deep ensemble: aleatoric part (U_alea) | Transformer | 0.460 [0.459; 0.461] | 0.874 [0.872; 0.876] | 0.037 [0.037; 0.037] |
| Deep ensemble: mutual information (MI) | GRU | 0.474 [0.467; 0.482] | 0.800 [0.794; 0.809] | 0.049 [0.046; 0.051] |
| Deep ensemble: mutual information (MI) | GRU + SN | 0.448 [0.443; 0.454] | 0.830 [0.823; 0.831] | 0.038 [0.037; 0.039] |
| Deep ensemble: mutual information (MI) | GRU + SN/res | 0.464 [0.460; 0.472] | 0.824 [0.818; 0.827] | 0.040 [0.039; 0.041] |
| Deep ensemble: mutual information (MI) | LSTM | 0.488 [0.485; 0.496] | 0.780 [0.767; 0.791] | 0.059 [0.056; 0.063] |
| Deep ensemble: mutual information (MI) | LSTM + SN | 0.452 [0.446; 0.457] | 0.835 [0.823; 0.840] | 0.045 [0.043; 0.047] |
| Deep ensemble: mutual information (MI) | LSTM + SN/res | 0.461 [0.451; 0.467] | 0.835 [0.821; 0.845] | 0.043 [0.039; 0.048] |
| Deep ensemble: mutual information (MI) | Transformer | 0.457 [0.455; 0.460] | 0.855 [0.852; 0.856] | 0.042 [0.042; 0.043] |
| Density: class-conditional Gaussian | GRU | 0.516 [0.514; 0.519] | 0.789 [0.784; 0.793] | 0.048 [0.045; 0.051] |

| | | | | |
|---|---|---|---|---|
| Density: class-conditional Gaussian | GRU + SN | 0.541 [0.540; 0.543] | 0.735 [0.732; 0.738] | 0.059 [0.057; 0.061] |
| Density: class-conditional Gaussian | GRU + SN/res | 0.534 [0.532; 0.536] | 0.747 [0.742; 0.752] | 0.057 [0.055; 0.058] |
| Density: class-conditional Gaussian | LSTM | 0.531 [0.524; 0.539] | 0.728 [0.712; 0.744] | 0.072 [0.065; 0.080] |
| Density: class-conditional Gaussian | LSTM + SN | 0.556 [0.554; 0.558] | 0.755 [0.749; 0.761] | 0.063 [0.059; 0.067] |
| Density: class-conditional Gaussian | LSTM + SN/res | 0.555 [0.554; 0.557] | 0.745 [0.740; 0.749] | 0.064 [0.061; 0.067] |
| Density: class-conditional Gaussian | Transformer | 0.512 [0.503; 0.520] | 0.753 [0.740; 0.766] | 0.065 [0.058; 0.072] |
| Density: shared covariance across classes | GRU | 0.514 [0.511; 0.516] | 0.793 [0.788; 0.798] | – |
| Density: shared covariance across classes | GRU + SN | 0.544 [0.543; 0.546] | 0.730 [0.727; 0.734] | – |
| Density: shared covariance across classes | GRU + SN/res | 0.536 [0.534; 0.538] | 0.743 [0.738; 0.748] | – |
| Density: shared covariance across classes | LSTM | 0.537 [0.529; 0.544] | 0.698 [0.679; 0.718] | – |
| Density: shared covariance across classes | LSTM + SN | 0.561 [0.559; 0.563] | 0.737 [0.729; 0.744] | – |
| Density: shared covariance across classes | LSTM + SN/res | 0.561 [0.559; 0.563] | 0.726 [0.721; 0.731] | – |
| Density: shared covariance across classes | Transformer | 0.509 [0.501; 0.517] | 0.757 [0.744; 0.770] | – |
| Density: Gaussian mixture per class | GRU | 0.505 [0.503; 0.506] | 0.780 [0.774; 0.786] | 0.049 [0.046; 0.053] |
| Density: Gaussian mixture per class | GRU + SN | 0.532 [0.531; 0.534] | 0.724 [0.721; 0.727] | 0.060 [0.058; 0.062] |
| Density: Gaussian mixture per class | GRU + SN/res | 0.523 [0.521; 0.525] | 0.742 [0.736; 0.748] | 0.056 [0.055; 0.058] |
| Density: Gaussian mixture per class | LSTM | 0.519 [0.513; 0.526] | 0.780 [0.770; 0.791] | 0.054 [0.049; 0.058] |
| Density: Gaussian mixture per class | LSTM + SN | 0.536 [0.535; 0.538] | 0.772 [0.768; 0.776] | 0.056 [0.054; 0.059] |
| Density: Gaussian mixture per class | LSTM + SN/res | 0.544 [0.542; 0.547] | 0.753 [0.749; 0.758] | 0.060 [0.057; 0.062] |
| Density: Gaussian mixture per class | Transformer | 0.505 [0.497; 0.513] | 0.727 [0.711; 0.743] | 0.073 [0.065; 0.082] |
| Density: L2-normalized representations | GRU | 0.495 [0.492; 0.498] | 0.844 [0.840; 0.848] | 0.038 [0.037; 0.038] |

| | | | | |
|---|---|---|---|---|
| Density: L2-normalized representations | GRU + SN | 0.491 [0.489; 0.494] | 0.833 [0.828; 0.838] | 0.039 [0.038; 0.039] |
| Density: L2-normalized representations | GRU + SN/res | 0.483 [0.482; 0.485] | 0.837 [0.831; 0.843] | 0.038 [0.037; 0.038] |
| Density: L2-normalized representations | LSTM | 0.500 [0.496; 0.505] | 0.832 [0.828; 0.836] | 0.045 [0.043; 0.047] |
| Density: L2-normalized representations | LSTM + SN | 0.501 [0.500; 0.503] | 0.852 [0.849; 0.855] | 0.040 [0.040; 0.041] |
| Density: L2-normalized representations | LSTM + SN/res | 0.497 [0.496; 0.499] | 0.850 [0.847; 0.853] | 0.040 [0.039; 0.040] |
| Density: L2-normalized representations | Transformer | 0.496 [0.488; 0.503] | 0.758 [0.745; 0.771] | 0.064 [0.057; 0.071] |
| Density: relative density score | GRU | 0.513 [0.511; 0.516] | 0.506 [0.489; 0.523] | 0.210 [0.206; 0.214] |
| Density: relative density score | GRU + SN | 0.492 [0.489; 0.495] | 0.555 [0.543; 0.567] | 0.174 [0.169; 0.179] |
| Density: relative density score | GRU + SN/res | 0.497 [0.494; 0.500] | 0.566 [0.557; 0.575] | 0.171 [0.166; 0.176] |
| Density: relative density score | LSTM | 0.500 [0.496; 0.504] | 0.676 [0.651; 0.701] | 0.155 [0.147; 0.162] |
| Density: relative density score | LSTM + SN | 0.450 [0.448; 0.451] | 0.676 [0.664; 0.689] | 0.160 [0.156; 0.163] |
| Density: relative density score | LSTM + SN/res | 0.456 [0.454; 0.458] | 0.721 [0.713; 0.729] | 0.131 [0.129; 0.134] |
| Density: relative density score | Transformer | 0.514 [0.511; 0.517] | 0.489 [0.469; 0.509] | 0.250 [0.239; 0.260] |
| Density: relative Mahalanobis distance (Ren et al.) | GRU | 0.519 [0.515; 0.523] | 0.555 [0.528; 0.582] | 0.198 [0.196; 0.201] |
| Density: relative Mahalanobis distance (Ren et al.) | GRU + SN | 0.507 [0.504; 0.510] | 0.484 [0.459; 0.508] | 0.209 [0.206; 0.211] |
| Density: relative Mahalanobis distance (Ren et al.) | GRU + SN/res | 0.500 [0.497; 0.503] | 0.516 [0.498; 0.533] | 0.197 [0.193; 0.200] |
| Density: relative Mahalanobis distance (Ren et al.) | LSTM | 0.517 [0.515; 0.519] | 0.595 [0.572; 0.619] | 0.196 [0.193; 0.198] |
| Density: relative Mahalanobis distance (Ren et al.) | LSTM + SN | 0.507 [0.505; 0.509] | 0.543 [0.523; 0.563] | 0.220 [0.219; 0.221] |
| Density: relative Mahalanobis distance (Ren et al.) | LSTM + SN/res | 0.499 [0.496; 0.502] | 0.548 [0.524; 0.571] | 0.212 [0.209; 0.215] |
| Density: relative Mahalanobis distance (Ren et al.) | Transformer | 0.512 [0.509; 0.515] | 0.488 [0.445; 0.531] | 0.252 [0.247; 0.257] |

Cohort separation: AUROC for external versus internal patients (0.5 = chance). Error detection: AUROC for misclassified patients on the external cohort at the operating threshold 0.1288. Selective prediction: excess area under the risk–coverage curve on the external cohort (lower is better). Values are means over ten seed models with the t-interval across paired seeds in brackets; for deep ensembles, the value of the full ensemble with the range over the ten nine-member ensembles. Monte Carlo dropout measures are available for the three base architectures. SN: spectral normalization; res: residual block.

**Supplementary Table 7 | Rank correlation between density measures and the predictive entropy of the same model on the external cohort**

| Model configuration | Density on L2-normalized representations | Class-conditional Gaussian density |
|---|---|---|
| **GRU** | 0.917 [0.89; 0.93] | 0.794 [0.72; 0.84] |
| **GRU + SN** | 0.768 [0.75; 0.79] | 0.439 [0.40; 0.48] |
| **GRU + SN/res** | 0.795 [0.78; 0.82] | 0.447 [0.42; 0.51] |
| **LSTM** | 0.643 [0.58; 0.73] | 0.314 [0.23; 0.40] |
| **LSTM + SN** | 0.738 [0.72; 0.75] | 0.456 [0.42; 0.49] |
| **LSTM + SN/res** | 0.747 [0.73; 0.77] | 0.412 [0.39; 0.44] |
| **Transformer** | 0.731 [0.65; 0.80] | 0.715 [0.63; 0.79] |

Spearman rank correlation per patient on the external cohort, mean over ten seed models with the range over seeds in brackets. The decision rule was stated before computation: $|\rho| \geq 0.70$ indicates a measure that largely tracks the prediction, $|\rho| < 0.40$ a measure that is largely independent of it. SN: spectral normalization; res: residual block.

**Supplementary Table 8 | Sensitivity of error detection to the operating threshold**

| Uncertainty measure | Configuration | Error detection, $\tau = 0.1288$ (main analysis) | Error detection, $\tau = 0.5$ |
|---|---|---|---|
| **Single model: predictive entropy** | GRU | 0.856 [0.849; 0.863] | 0.755 [0.752; 0.758] |
| **Single model: predictive entropy** | GRU + SN | 0.845 [0.837; 0.852] | 0.760 [0.758; 0.762] |
| **Single model: predictive entropy** | GRU + SN/res | 0.844 [0.837; 0.850] | 0.759 [0.757; 0.761] |
| **Single model: predictive entropy** | LSTM | 0.858 [0.853; 0.864] | 0.744 [0.740; 0.748] |
| **Single model: predictive entropy** | LSTM + SN | 0.863 [0.858; 0.868] | 0.756 [0.753; 0.758] |
| **Single model: predictive entropy** | LSTM + SN/res | 0.859 [0.854; 0.863] | 0.755 [0.752; 0.757] |
| **Single model: predictive entropy** | Transformer | 0.870 [0.861; 0.880] | 0.767 [0.762; 0.772] |
| **MC dropout: predictive entropy (U_total)** | GRU | 0.857 [0.851; 0.863] | 0.761 [0.759; 0.763] |
| **MC dropout: predictive entropy (U_total)** | LSTM | 0.861 [0.856; 0.866] | 0.753 [0.749; 0.756] |
| **MC dropout: predictive entropy (U_total)** | Transformer | 0.869 [0.860; 0.879] | 0.770 [0.764; 0.775] |
| **MC dropout: aleatoric part (U_alea)** | GRU | 0.858 [0.851; 0.864] | 0.760 [0.758; 0.762] |
| **MC dropout: aleatoric part (U_alea)** | LSTM | 0.861 [0.856; 0.866] | 0.751 [0.748; 0.755] |
| **MC dropout: aleatoric part (U_alea)** | Transformer | 0.869 [0.860; 0.879] | 0.770 [0.764; 0.775] |
| **MC dropout: mutual information (MI)** | GRU | 0.810 [0.806; 0.814] | 0.742 [0.739; 0.745] |
| **MC dropout: mutual information (MI)** | LSTM | 0.791 [0.780; 0.801] | 0.731 [0.725; 0.737] |
| **MC dropout: mutual information (MI)** | Transformer | 0.836 [0.828; 0.843] | 0.738 [0.734; 0.743] |
| **MC dropout: logit variance (epistemic)** | GRU | 0.326 [0.313; 0.339] | 0.418 [0.409; 0.427] |
| **MC dropout: logit variance (epistemic)** | LSTM | 0.314 [0.296; 0.333] | 0.427 [0.414; 0.439] |
| **MC dropout: logit variance (epistemic)** | Transformer | 0.181 [0.169; 0.193] | 0.278 [0.270; 0.286] |
| **Deep ensemble: predictive entropy (U_total)** | GRU | 0.858 [0.856; 0.860] | 0.757 [0.755; 0.757] |
| **Deep ensemble: predictive entropy (U_total)** | GRU + SN | 0.846 [0.844; 0.848] | 0.761 [0.761; 0.762] |
| **Deep ensemble: predictive entropy (U_total)** | GRU + SN/res | 0.845 [0.843; 0.847] | 0.762 [0.761; 0.762] |
| **Deep ensemble: predictive entropy (U_total)** | LSTM | 0.861 [0.859; 0.862] | 0.748 [0.747; 0.749] |

| | | | |
|---|---|---|---|
| **Deep ensemble: predictive entropy (U_total)** | LSTM + SN | 0.864 [0.862; 0.865] | 0.756 [0.755; 0.756] |
| **Deep ensemble: predictive entropy (U_total)** | LSTM + SN/res | 0.859 [0.858; 0.860] | 0.755 [0.754; 0.755] |
| **Deep ensemble: predictive entropy (U_total)** | Transformer | 0.874 [0.872; 0.876] | 0.767 [0.766; 0.768] |
| **Deep ensemble: aleatoric part (U_alea)** | GRU | 0.858 [0.856; 0.860] | 0.756 [0.755; 0.757] |
| **Deep ensemble: aleatoric part (U_alea)** | GRU + SN | 0.846 [0.844; 0.848] | 0.761 [0.760; 0.762] |
| **Deep ensemble: aleatoric part (U_alea)** | GRU + SN/res | 0.845 [0.843; 0.847] | 0.761 [0.761; 0.762] |
| **Deep ensemble: aleatoric part (U_alea)** | LSTM | 0.861 [0.859; 0.862] | 0.747 [0.746; 0.748] |
| **Deep ensemble: aleatoric part (U_alea)** | LSTM + SN | 0.864 [0.862; 0.865] | 0.756 [0.755; 0.756] |
| **Deep ensemble: aleatoric part (U_alea)** | LSTM + SN/res | 0.859 [0.858; 0.860] | 0.754 [0.754; 0.755] |
| **Deep ensemble: aleatoric part (U_alea)** | Transformer | 0.874 [0.872; 0.876] | 0.767 [0.766; 0.768] |
| **Deep ensemble: mutual information (MI)** | GRU | 0.800 [0.794; 0.809] | 0.735 [0.730; 0.741] |
| **Deep ensemble: mutual information (MI)** | GRU + SN | 0.830 [0.823; 0.831] | 0.751 [0.746; 0.753] |
| **Deep ensemble: mutual information (MI)** | GRU + SN/res | 0.824 [0.818; 0.827] | 0.745 [0.740; 0.748] |
| **Deep ensemble: mutual information (MI)** | LSTM | 0.780 [0.767; 0.791] | 0.711 [0.704; 0.719] |
| **Deep ensemble: mutual information (MI)** | LSTM + SN | 0.835 [0.823; 0.840] | 0.728 [0.719; 0.733] |
| **Deep ensemble: mutual information (MI)** | LSTM + SN/res | 0.835 [0.821; 0.845] | 0.741 [0.726; 0.754] |
| **Deep ensemble: mutual information (MI)** | Transformer | 0.855 [0.852; 0.856] | 0.746 [0.743; 0.748] |
| **Density: class-conditional Gaussian** | GRU | 0.789 [0.784; 0.793] | 0.738 [0.734; 0.742] |
| **Density: class-conditional Gaussian** | GRU + SN | 0.735 [0.732; 0.738] | 0.697 [0.694; 0.700] |
| **Density: class-conditional Gaussian** | GRU + SN/res | 0.747 [0.742; 0.752] | 0.700 [0.699; 0.702] |
| **Density: class-conditional Gaussian** | LSTM | 0.728 [0.712; 0.744] | 0.699 [0.692; 0.705] |
| **Density: class-conditional Gaussian** | LSTM + SN | 0.755 [0.749; 0.761] | 0.708 [0.706; 0.710] |
| **Density: class-conditional Gaussian** | LSTM + SN/res | 0.745 [0.740; 0.749] | 0.704 [0.702; 0.707] |
| **Density: class-conditional Gaussian** | Transformer | 0.753 [0.740; 0.766] | 0.699 [0.688; 0.710] |

| | | | |
|---|---|---|---|
| **Density: Gaussian mixture per class** | GRU | 0.780 [0.774; 0.786] | 0.739 [0.734; 0.743] |
| **Density: Gaussian mixture per class** | GRU + SN | 0.724 [0.721; 0.727] | 0.692 [0.689; 0.695] |
| **Density: Gaussian mixture per class** | GRU + SN/res | 0.742 [0.736; 0.748] | 0.700 [0.698; 0.703] |
| **Density: Gaussian mixture per class** | LSTM | 0.780 [0.770; 0.791] | 0.726 [0.720; 0.731] |
| **Density: Gaussian mixture per class** | LSTM + SN | 0.772 [0.768; 0.776] | 0.720 [0.718; 0.721] |
| **Density: Gaussian mixture per class** | LSTM + SN/res | 0.753 [0.749; 0.758] | 0.711 [0.708; 0.714] |
| **Density: Gaussian mixture per class** | Transformer | 0.727 [0.711; 0.743] | 0.688 [0.676; 0.701] |
| **Density: L2-normalized representations** | GRU | 0.844 [0.840; 0.848] | 0.764 [0.762; 0.766] |
| **Density: L2-normalized representations** | GRU + SN | 0.833 [0.828; 0.838] | 0.758 [0.756; 0.759] |
| **Density: L2-normalized representations** | GRU + SN/res | 0.837 [0.831; 0.843] | 0.758 [0.756; 0.760] |
| **Density: L2-normalized representations** | LSTM | 0.832 [0.828; 0.836] | 0.751 [0.748; 0.754] |
| **Density: L2-normalized representations** | LSTM + SN | 0.852 [0.849; 0.855] | 0.758 [0.756; 0.761] |
| **Density: L2-normalized representations** | LSTM + SN/res | 0.850 [0.847; 0.853] | 0.759 [0.757; 0.760] |
| **Density: L2-normalized representations** | Transformer | 0.758 [0.745; 0.771] | 0.704 [0.693; 0.715] |
| **Density: relative density score** | GRU | 0.506 [0.489; 0.523] | 0.460 [0.456; 0.464] |
| **Density: relative density score** | GRU + SN | 0.555 [0.543; 0.567] | 0.497 [0.493; 0.501] |
| **Density: relative density score** | GRU + SN/res | 0.566 [0.557; 0.575] | 0.502 [0.499; 0.506] |
| **Density: relative density score** | LSTM | 0.676 [0.651; 0.701] | 0.550 [0.538; 0.563] |
| **Density: relative density score** | LSTM + SN | 0.676 [0.664; 0.689] | 0.526 [0.521; 0.531] |
| **Density: relative density score** | LSTM + SN/res | 0.721 [0.713; 0.729] | 0.569 [0.562; 0.576] |
| **Density: relative density score** | Transformer | 0.489 [0.469; 0.509] | 0.430 [0.420; 0.440] |

Means over ten seed models with the 95% t-interval across seeds in brackets; for deep ensembles, the value of the full ensemble with the range over the ten nine-member ensembles. τ: classification threshold. SN: spectral normalization; res: residual block.

**Supplementary Table 9 | Misclassifications captured and missed when the 20% most uncertain patients are deferred by predictive entropy, external cohort**

| Quantity | GRU | LSTM | Transformer |
|---|---|---|---|
| **Misclassified predictions, all patients** | 17.5% | 18.9% | 20.1% |
| **Missed deaths among all misclassifications** | 30.1% [23.6; 37.2] | 26.2% [20.1; 30.9] | 24.9% [20.0; 40.2] |
| **False-positive predictions captured by deferral** | 98.7% [94.1; 99.9] | 96.0% [81.8; 99.2] | 90.5% [81.5; 100.0] |
| **Missed deaths captured by deferral** | 7.0% [0.0; 14.8] | 3.4% [0.0; 9.1] | 2.4% [0.0; 17.3] |
| **Misclassified among patients not deferred** | 6.3% | 6.7% | 8.0% |
| **Missed deaths among misclassifications not deferred** | 96.5% [84.0; 99.8] | 90.0% [58.0; 97.7] | 77.5% [58.0; 100.0] |
| **Predicted risk of missed deaths, median** | 3.6% [3.1; 4.3] | 3.5% [2.8; 4.0] | 3.2% [2.6; 3.6] |
| **Missed deaths with predicted risk below 5%** | 60.7% [55.0; 64.2] | 61.9% [58.3; 66.6] | 65.5% [61.5; 70.5] |

Means over ten seed models with the range over seeds in brackets. Misclassified: flagged at risk (predicted probability above 0.1288) and survived (false-positive prediction), or not flagged and died (missed death). Deferral: the 20% of patients with the highest predictive entropy of the single model; the remaining 80% are not deferred.

**Supplementary Table 10 | Validation loss at the end of training and at its minimum, by architecture and variant**

| Configuration | Epochs | Validation loss, last epoch | Validation loss, minimum | Relative increase, mean (max) | Epoch of minimum, median | Seeds with minimum at last epoch |
|---|---|---|---|---|---|---|
| GRU | 28 | 0.2767 | 0.2705 | 2.3% (7.6%) | 23 | 1/10 |
| GRU + SN | 28 | 0.2824 | 0.2779 | 1.6% (4.8%) | 23 | 2/10 |
| GRU + SN/res | 28 | 0.2815 | 0.2776 | 1.4% (4.3%) | 23 | 0/10 |
| LSTM | 40 | 0.2755 | 0.2709 | 1.7% (2.8%) | 29 | 0/10 |
| LSTM + SN | 40 | 0.2770 | 0.2757 | 0.5% (1.3%) | 36 | 0/10 |
| LSTM + SN/res | 40 | 0.2770 | 0.2753 | 0.6% (1.5%) | 35 | 1/10 |
| Transformer | 13 | 0.2862 | 0.2819 | 1.5% (5.7%) | 9 | 0/10 |

Means over ten seed models. Minimum: each seed model's own lowest validation loss. Relative increase: last epoch above that minimum, mean (largest) over seeds. SN: spectral normalization; res: residual block.

**Supplementary Table 11 | Sensitivity analysis: models taken at the epoch of their lowest validation loss versus models at the end of training, three base architectures**

| | End of training (as reported) | Epoch of lowest validation loss | Paired difference [95% interval] | Largest single-seed difference | Seed interval half-width | Within seed variation |
|---|---|---|---|---|---|---|
| **GRU** | | | | | | |
| AUROC, internal test set | 0.8732 | 0.8753 | +0.0021 [-0.0002; +0.0044] | 0.0079 | 0.0026 | yes |
| AUROC, external cohort | 0.7732 | 0.7772 | +0.0041 [+0.0007; +0.0074] | 0.0100 | 0.0027 | no |
| Calibration slope, internal test set | 0.962 | 1.014 | +0.052 [+0.021; +0.083] | 0.126 | 0.027 | no |
| Calibration slope, external cohort | 0.645 | 0.682 | +0.037 [+0.019; +0.056] | 0.075 | 0.014 | no |
| Cross-entropy, internal test set | 0.2836 | 0.2781 | -0.0055 [-0.0105; -0.0006] | 0.0205 | 0.0054 | no |
| Cross-entropy, external cohort | 0.3084 | 0.3013 | -0.0071 [-0.0107; -0.0036] | 0.0146 | 0.0042 | no |
| Predictive entropy: cohort separation (AUROC) | 0.4749 | 0.4642 | -0.0107 [-0.0175; -0.0038] | 0.0241 | 0.0044 | no |
| Predictive entropy: error detection (AUROC) | 0.8559 | 0.8613 | +0.0054 [-0.0006; +0.0114] | 0.0195 | 0.0069 | yes |
| Predictive entropy: E-AURC | 0.0378 | 0.0372 | -0.0006 [-0.0016; +0.0005] | 0.0024 | 0.0006 | yes |
| Error rate at 80% coverage | 0.0627 | 0.0611 | -0.0016 [-0.0028; -0.0005] | 0.0058 | 0.0023 | yes |
| Error rate at the threshold | 0.1749 | 0.1804 | +0.0054 [-0.0026; +0.0135] | 0.0252 | 0.0098 | yes |
| **LSTM** | | | | | | |
| AUROC, internal test set | 0.8678 | 0.8713 | +0.0035 [+0.0019; +0.0051] | 0.0063 | 0.0017 | no |
| AUROC, external cohort | 0.7623 | 0.7680 | +0.0057 [+0.0021; +0.0092] | 0.0133 | 0.0037 | no |
| Calibration slope, internal test set | 0.915 | 1.023 | +0.108 [+0.037; +0.179] | 0.300 | 0.049 | no |
| Calibration slope, external cohort | 0.569 | 0.638 | +0.069 [+0.029; +0.109] | 0.182 | 0.030 | no |
| Cross-entropy, internal test set | 0.2829 | 0.2769 | -0.0060 [-0.0082; -0.0038] | 0.0096 | 0.0020 | no |

| | End of training (as reported) | Epoch of lowest validation loss | Paired difference [95% interval] | Largest single-seed difference | Seed interval half-width | Within seed variation |
|---|---|---|---|---|---|---|
| Cross-entropy, external cohort | 0.3172 | 0.3051 | -0.0121 [-0.0185; -0.0056] | 0.0313 | 0.0050 | no |
| Predictive entropy: cohort separation (AUROC) | 0.4658 | 0.4520 | -0.0138 [-0.0221; -0.0055] | 0.0371 | 0.0048 | no |
| Predictive entropy: error detection (AUROC) | 0.8583 | 0.8653 | +0.0070 [-0.0009; +0.0150] | 0.0187 | 0.0053 | no |
| Predictive entropy: E-AURC | 0.0422 | 0.0405 | -0.0016 [-0.0030; -0.0003] | 0.0042 | 0.0013 | no |
| Error rate at 80% coverage | 0.0672 | 0.0750 | +0.0078 [-0.0062; +0.0217] | 0.0345 | 0.0077 | no |
| Error rate at the threshold | 0.1891 | 0.1993 | +0.0102 [-0.0066; +0.0270] | 0.0457 | 0.0108 | yes |
| **Transformer** | | | | | | |
| AUROC, internal test set | 0.8581 | 0.8570 | -0.0011 [-0.0024; +0.0001] | 0.0034 | 0.0015 | yes |
| AUROC, external cohort | 0.7658 | 0.7663 | +0.0005 [-0.0028; +0.0038] | 0.0083 | 0.0016 | yes |
| Calibration slope, internal test set | 0.843 | 0.917 | +0.075 [+0.031; +0.118] | 0.227 | 0.031 | no |
| Calibration slope, external cohort | 0.593 | 0.637 | +0.044 [+0.015; +0.072] | 0.139 | 0.020 | no |
| Cross-entropy, internal test set | 0.2969 | 0.2925 | -0.0044 [-0.0102; +0.0015] | 0.0255 | 0.0052 | yes |
| Cross-entropy, external cohort | 0.3163 | 0.3087 | -0.0077 [-0.0119; -0.0035] | 0.0185 | 0.0036 | no |
| Predictive entropy: cohort separation (AUROC) | 0.4617 | 0.4540 | -0.0077 [-0.0123; -0.0031] | 0.0229 | 0.0035 | no |
| Predictive entropy: error detection (AUROC) | 0.8704 | 0.8728 | +0.0024 [-0.0089; +0.0136] | 0.0371 | 0.0098 | yes |
| Predictive entropy: E-AURC | 0.0373 | 0.0372 | -0.0001 [-0.0011; +0.0009] | 0.0022 | 0.0005 | yes |
| Error rate at 80% coverage | 0.0796 | 0.0805 | +0.0008 [-0.0134; +0.0150] | 0.0325 | 0.0099 | yes |
| Error rate at the threshold | 0.2014 | 0.2062 | +0.0048 [-0.0140; +0.0237] | 0.0555 | 0.0159 | yes |

Means over ten seed models; paired difference: epoch of lowest validation loss minus end of training, per seed, with the 95% t-interval. Seed interval half-width: half-width of the 95% t-interval of the end-of-training value. Within seed variation: yes if the absolute paired difference does not exceed this half-width.

**Supplementary Table 12 | Error detection and selective prediction on the internal test set and the external cohort, three base architectures**

| Uncertainty measure | Model family | Error detection (AUROC), internal test set | Error detection (AUROC), external cohort | Error detection, paired difference external − internal [95% interval] | E-AURC, internal test set | E-AURC, external cohort | E-AURC, paired difference external − internal [95% interval] | Risk at 80% coverage, internal test set | Risk at 80% coverage, external cohort |
|---|---|---|---|---|---|---|---|---|---|
| Single model: predictive entropy | GRU | 0.873 [0.870; 0.876] | 0.856 [0.849; 0.863] | -0.017 [-0.022; -0.013] | 0.026 [0.025; 0.027] | 0.038 [0.037; 0.038] | 0.012 [0.011; 0.013] | 0.087 [0.072; 0.101] | 0.063 [0.060; 0.065] |
| Single model: predictive entropy | LSTM | 0.878 [0.876; 0.881] | 0.858 [0.853; 0.864] | -0.020 [-0.024; -0.016] | 0.028 [0.027; 0.029] | 0.042 [0.041; 0.043] | 0.014 [0.013; 0.015] | 0.104 [0.090; 0.118] | 0.067 [0.060; 0.075] |
| Single model: predictive entropy | Transformer | 0.878 [0.871; 0.884] | 0.870 [0.861; 0.880] | -0.007 [-0.011; -0.004] | 0.028 [0.027; 0.029] | 0.037 [0.037; 0.038] | 0.010 [0.009; 0.010] | 0.107 [0.090; 0.125] | 0.080 [0.070; 0.090] |
| MC dropout: predictive entropy (U_total) | GRU | 0.877 [0.874; 0.880] | 0.857 [0.851; 0.863] | -0.020 [-0.023; -0.016] | 0.025 [0.024; 0.026] | 0.037 [0.036; 0.037] | 0.012 [0.011; 0.013] | 0.088 [0.077; 0.100] | 0.067 [0.062; 0.072] |
| MC dropout: predictive entropy (U_total) | LSTM | 0.879 [0.877; 0.882] | 0.861 [0.856; 0.866] | -0.018 [-0.023; -0.014] | 0.027 [0.026; 0.028] | 0.040 [0.039; 0.042] | 0.013 [0.012; 0.014] | 0.106 [0.092; 0.120] | 0.071 [0.063; 0.080] |
| MC dropout: predictive entropy (U_total) | Transformer | 0.875 [0.869; 0.881] | 0.869 [0.860; 0.879] | -0.005 [-0.009; -0.001] | 0.029 [0.028; 0.030] | 0.038 [0.037; 0.038] | 0.009 [0.008; 0.010] | 0.110 [0.091; 0.128] | 0.081 [0.070; 0.091] |
| MC dropout: mutual information (MI) | GRU | 0.826 [0.822; 0.829] | 0.810 [0.806; 0.814] | -0.015 [-0.019; -0.012] | 0.037 [0.034; 0.039] | 0.047 [0.045; 0.048] | 0.010 [0.009; 0.011] | 0.113 [0.103; 0.124] | 0.103 [0.095; 0.111] |
| MC dropout: mutual information (MI) | LSTM | 0.804 [0.791; 0.817] | 0.791 [0.780; 0.801] | -0.013 [-0.018; -0.008] | 0.047 [0.041; 0.053] | 0.056 [0.051; 0.061] | 0.009 [0.007; 0.011] | 0.133 [0.120; 0.147] | 0.120 [0.108; 0.131] |
| MC dropout: mutual information (MI) | Transformer | 0.845 [0.840; 0.850] | 0.836 [0.828; 0.843] | -0.009 [-0.014; -0.005] | 0.038 [0.036; 0.041] | 0.047 [0.046; 0.049] | 0.009 [0.007; 0.011] | 0.121 [0.106; 0.136] | 0.109 [0.096; 0.121] |

| | | | | | | | | | |
|---|---|---|---|---|---|---|---|---|---|
| MC dropout: logit variance (epistemic) | GRU | 0.290 [0.276; 0.305] | 0.326 [0.313; 0.339] | 0.036 [0.032; 0.040] | 0.284 [0.266; 0.302] | 0.273 [0.258; 0.289] | -0.011 [-0.015; -0.006] | 0.204 [0.189; 0.219] | 0.190 [0.178; 0.202] |
| MC dropout: logit variance (epistemic) | LSTM | 0.266 [0.250; 0.283] | 0.314 [0.296; 0.333] | 0.048 [0.042; 0.054] | 0.316 [0.296; 0.336] | 0.298 [0.279; 0.317] | -0.018 [-0.023; -0.013] | 0.229 [0.214; 0.245] | 0.205 [0.192; 0.219] |
| MC dropout: logit variance (epistemic) | Transformer | 0.176 [0.165; 0.186] | 0.181 [0.169; 0.193] | 0.006 [0.001; 0.011] | 0.353 [0.325; 0.381] | 0.365 [0.340; 0.391] | 0.012 [0.007; 0.018] | 0.249 [0.227; 0.272] | 0.242 [0.222; 0.262] |
| Deep ensemble: predictive entropy (U_total) | GRU | 0.875 [0.874; 0.875] | 0.858 [0.856; 0.860] | -0.016 [-0.018; -0.016] | 0.026 [0.025; 0.026] | 0.037 [0.037; 0.037] | 0.012 [0.011; 0.012] | 0.090 [0.087; 0.093] | 0.060 [0.060; 0.061] |
| Deep ensemble: predictive entropy (U_total) | LSTM | 0.879 [0.879; 0.880] | 0.861 [0.859; 0.862] | -0.018 [-0.020; -0.017] | 0.027 [0.027; 0.027] | 0.041 [0.041; 0.041] | 0.014 [0.014; 0.014] | 0.104 [0.101; 0.107] | 0.061 [0.061; 0.061] |
| Deep ensemble: predictive entropy (U_total) | Transformer | 0.877 [0.876; 0.878] | 0.874 [0.872; 0.876] | -0.003 [-0.004; -0.002] | 0.028 [0.028; 0.028] | 0.037 [0.037; 0.037] | 0.009 [0.009; 0.009] | 0.118 [0.112; 0.125] | 0.080 [0.077; 0.088] |
| Deep ensemble: mutual information (MI) | GRU | 0.813 [0.809; 0.820] | 0.800 [0.794; 0.809] | -0.013 [-0.016; -0.011] | 0.040 [0.038; 0.041] | 0.049 [0.046; 0.051] | 0.009 [0.008; 0.010] | 0.120 [0.116; 0.121] | 0.105 [0.102; 0.109] |
| Deep ensemble: mutual information (MI) | LSTM | 0.799 [0.787; 0.809] | 0.780 [0.767; 0.791] | -0.019 [-0.024; -0.018] | 0.047 [0.045; 0.051] | 0.059 [0.056; 0.063] | 0.012 [0.011; 0.014] | 0.141 [0.137; 0.144] | 0.125 [0.119; 0.129] |
| Deep ensemble: mutual information (MI) | Transformer | 0.865 [0.860; 0.865] | 0.855 [0.852; 0.856] | -0.010 [-0.011; -0.007] | 0.032 [0.031; 0.034] | 0.042 [0.042; 0.043] | 0.011 [0.009; 0.011] | 0.117 [0.114; 0.125] | 0.104 [0.102; 0.111] |
| Density: class-conditional Gaussian | GRU | 0.800 [0.794; 0.806] | 0.789 [0.784; 0.793] | -0.011 [-0.013; -0.009] | 0.042 [0.038; 0.045] | 0.048 [0.045; 0.051] | 0.006 [0.005; 0.007] | 0.129 [0.117; 0.140] | 0.120 [0.110; 0.130] |
| Density: class-conditional Gaussian | LSTM | 0.728 [0.712; 0.744] | 0.728 [0.712; 0.744] | 0.000 [-0.006; 0.007] | 0.073 [0.065; 0.081] | 0.072 [0.065; 0.080] | -0.001 [-0.003; 0.002] | 0.155 [0.141; 0.169] | 0.144 [0.130; 0.158] |
| Density: class-conditional Gaussian | Transformer | 0.755 [0.742; 0.768] | 0.753 [0.740; 0.766] | -0.003 [-0.009; 0.003] | 0.061 [0.054; 0.069] | 0.065 [0.058; 0.072] | 0.004 [0.002; 0.006] | 0.156 [0.138; 0.174] | 0.152 [0.137; 0.167] |

Means over ten seed models with the 95% t-interval across seeds in brackets; differences are paired per seed. For deep ensembles, the value of the full ensemble with the range over the ten nine-member ensembles. Error detection: AUROC for misclassified patients at the operating threshold 0.1288. E-AURC: excess area under the risk–coverage curve (lower is better). Risk at 80% coverage: misclassification rate among the accepted patients when the 20% most uncertain are deferred; not directly comparable between cohorts, because the fixed classification threshold yields a different mix of false alarms and missed deaths (Supplementary Note 8). Internal test set: 7,298 stays; external cohort: 66,934 stays.

### References


1. Liu JZ, Lin Z, Padhy S, Tran D, Bedrax-Weiss T, Lakshminarayanan B. Simple and Principled Uncertainty Estimation with Deterministic Deep Learning via Distance Awareness. *arXiv*. Preprint posted online October 26, 2020:arXiv:2006.10108. doi:10.48550/arXiv.2006.10108

2. Mukhoti J, Kirsch A, Amersfoort J van, Torr PHS, Gal Y. Deep Deterministic Uncertainty: A Simple Baseline. *arXiv*. Preprint posted online January 28, 2022:arXiv:2102.11582. doi:10.48550/arXiv.2102.11582

3. Akiba T, Sano S, Yanase T, Ohta T, Koyama M. Optuna: A Next-generation Hyperparameter Optimization Framework. *arXiv*. Preprint posted online July 25, 2019:arXiv:1907.10902. doi:10.48550/arXiv.1907.10902

4. Bergstra JS, Bardenet R, Bengio Y, Kégl B. Algorithms for Hyper-Parameter Optimization. In: *Advances in Neural Information Processing Systems 24*. Curran Associates; 2011:2546-2554. https://papers.nips.cc/paper/2011/hash/86e8f7ab32cfd12577bc2619bc635690-Abstract.html

5. Pakdaman Naeini M, Cooper G, Hauskrecht M. Obtaining Well Calibrated Probabilities Using Bayesian Binning. *Proc AAAI Conf Artif Intell*. 2015;29(1). doi:10.1609/aaai.v29i1.9602

6. Guo C, Pleiss G, Sun Y, Weinberger KQ. On Calibration of Modern Neural Networks. *arXiv*. Preprint posted online August 3, 2017:arXiv:1706.04599. doi:10.48550/arXiv.1706.04599

7. Nixon J, Dusenberry M, Jerfel G, et al. Measuring Calibration in Deep Learning. *arXiv*. Preprint posted online August 7, 2020:arXiv:1904.01685. doi:10.48550/arXiv.1904.01685

8. Sun Y, Ming Y, Zhu X, Li Y. Out-of-Distribution Detection with Deep Nearest Neighbors. *arXiv*. Preprint posted online December 8, 2022:arXiv:2204.06507. doi:10.48550/arXiv.2204.06507

9. Ren J, Fort S, Liu J, Roy AG, Padhy S, Lakshminarayanan B. A Simple Fix to Mahalanobis Distance for Improving Near-OOD Detection. *arXiv*. Preprint posted online June 16, 2021:arXiv:2106.09022. doi:10.48550/arXiv.2106.09022

10. Prechelt L. Early stopping — but when? In: Montavon G, Orr GB, Müller KR, eds. *Neural Networks: Tricks of the Trade: Second Edition*. Springer Berlin Heidelberg; 2012:53-67. doi:10.1007/978-3-642-35289-8_5

11. Lodwich A, Rangoni Y, Breuel T. Evaluation of robustness and performance of Early Stopping Rules with Multi Layer Perceptrons. In: *2009 International Joint Conference on Neural Networks*. IEEE; 2009:1877-1884. doi:10.1109/IJCNN.2009.5178626

12. Harutyunyan H, Khachatrian H, Kale DC, Ver Steeg G, Galstyan A. Multitask learning and benchmarking with clinical time series data. *Sci Data*. 2019;6(1):96. doi:10.1038/s41597-019-0103-9
